# Disrupted Limbic-Prefrontal Effective Connectivity in Response to Fearful Faces in Lifetime Depression

**DOI:** 10.1101/2023.07.18.23292846

**Authors:** Aleks Stolicyn, Mathew A. Harris, Laura de Nooij, Xueyi Shen, Jennifer A. Macfarlane, Archie Campbell, Christopher J. McNeil, Anca-Larisa Sandu, Alison D. Murray, Gordon D. Waiter, Stephen M. Lawrie, Douglas J. Steele, Andrew M. McIntosh, Liana Romaniuk, Heather C. Whalley

**Affiliations:** Division of Psychiatry, Centre for Clinical Brain Sciences, University of Edinburgh, Kennedy Tower, Royal Edinburgh Hospital, Edinburgh EH10 5HF, United Kingdom; Donders Institute for Brain, Cognition and Behaviour, Radboud University Medical Center, Nijmegen 6525 EN, Netherlands; Division of Imaging Science and Technology, School of Medicine, University of Dundee, Dundee DD1 9SY, United Kingdom; Department of Medical Physics, NHS Tayside, Dundee DD2 1UB, United Kingdom; SINASPE Consortium, https://www.sinapse.ac.uk; Centre for Genomic and Experimental Medicine, Institute of Genetics and Cancer, University of Edinburgh, Edinburgh EH4 2XU, United Kingdom; Aberdeen Biomedical Imaging Centre, Institute of Medical Sciences, University of Aberdeen, Aberdeen AB25 2ZN, United Kingdom

**Keywords:** Major depressive disorder, amygdala, prefrontal cortex, fearful faces, functional MRI, effective connectivity

## Abstract

**Background:** Multiple brain imaging studies of negative emotional bias in major depressive disorder (MDD) have used images of fearful facial expressions and focused on the amygdala and the prefrontal cortex. The results have, however, been inconsistent, potentially due to small sample sizes (typically *N* < 50). It remains unclear if any alterations are a characteristic of current depression or of past experience of depression, and whether there are MDD-related changes in effective connectivity between the two brain regions.

**Methods:** Activations and effective connectivity between the amygdala and dorsolateral prefrontal cortex (DLPFC) in response to fearful face stimuli were studied in a large population-based sample from Generation Scotland. Participants either had no history of MDD (*N* = 664 in activation analyses, *N* = 474 in connectivity analyses) or had a diagnosis of MDD during their lifetime (LMDD, *N* = 290 in activation analyses, *N* = 214 in connectivity analyses). The within-scanner task involved implicit facial emotion processing of neutral and fearful faces.

**Results:** Compared to controls, LMDD was associated with increased activations in left amygdala (*P_FWE_* = 0.031 *, k _E_* = 4) and left DLPFC (*P_FWE_* = 0.002 *, k _E_* = 33), increased mean bilateral amygdala activation (*β* = 0.0715 *, P* = 0.0314), and increased inhibition from left amygdala to left DLPFC, all in response to fearful faces contrasted to baseline. Results did not appear to be attributable to depressive illness severity or antidepressant medication status at scan time.

**Limitations:** Most studied participants had past rather than current depression, average severity of ongoing depression symptoms was low, and a substantial proportion of participants were receiving medication. The study was not longitudinal and the participants were only assessed a single time.

**Conclusions:** LMDD is associated with hyperactivity of the amygdala and DLPFC, and with stronger amygdala to DLPFC inhibitory connectivity, all in response to fearful faces, unrelated to depression severity at scan time. These results help reduce inconsistency in past literature and suggest disruption of ‘bottom-up’ limbic-prefrontal effective connectivity in depression.

## Introduction

Depression (major depressive disorder, MDD) is a prevalent condition which on average affects between 10% and 15% of the general population over their lifetime (Kessler and Bromet, 2013; Lim et al., 2018). Depression has significant social and economic impacts and has been estimated to be one of the leading causes of years lived with disability (GBD 2017 Disease and Injury Incidence and Prevalence Collaborators, 2018). According to the primary diagnostic manuals (DSM-5 and ICD-10), the core indicators of depression are low mood and loss of interest (anhedonia), accompanied by a range of other somatic and/or cognitive symptoms (American Psychiatric Association, 2013; World Health Organisation, 1993).

An important clinical aspect of depression is negative cognitive bias – a tendency to attend to, focus on, and remember negative emotional information (Everaert et al., 2012; Gotlib and Joormann, 2010). Because negative emotional information is closely related to induction of low mood, negative bias is considered to contribute to maintenance of low mood symptoms in depression (Beck and Bredemeier, 2016; Clark and Beck, 2010). Behavioural changes related to negative bias have been shown in studies with stimuli such as emotional words, images of affectively valenced scenes, and images of emotional facial expressions (de Nooij et al., 2022; Elliott et al., 2011; Gotlib and Joormann, 2010; Miskowiak and Carvalho, 2014). It is theorised that in ongoing depression at the neural level the negative bias is underpinned by hyperactive limbic subcortical structures – primarily the amygdala – and hypoactive frontal cortical areas such as the dorsolateral prefrontal cortex (DLPFC) (DeRubeis et al., 2008; Disner et al., 2011; Groenewold et al., 2013; Roiser and Sahakian, 2013). The amygdala is thought to have a crucial role in processing emotional faces and fear-related stimuli (Adolphs, 2008; Fusar-Poli et al., 2009; Phelps and LeDoux, 2005; Whalen et al., 2013), and amygdala hyperactivity could be responsible for the stronger focus on negative information. Altered activity of the DLPFC, on the other hand, could represent changes in cognitive control and lower inhibition of emotional processing in the amygdala. It remains unclear whether altered activations of these regions are a characteristic of depressed state or of experience of depression over the lifetime, and if these changes are related to disrupted limbic-prefrontal (‘bottom-up’), or prefrontal-limbic (‘top-down’) effective connectivity. These questions are important to address because they can indicate which neural abnormality (amygdala or DLPFC) may be primary and causing downstream brain activation changes in depression, and if this is related to ongoing symptoms.

Over the past 15 years many functional brain imaging studies have used images of emotional facial expressions (fearful, sad, angry) to study neural mechanisms of negative bias in depression, with fearful faces most commonly used. The brain regions of interest (ROI) most frequently investigated in these studies were the amygdala and the prefrontal cortex (Supplementary Section S1 and Table S1). Despite the strong focus on the amygdala, only a subset of studies found increased activation in this region (Fales et al., 2008; Greening et al., 2013; Korgaonkar et al., 2019; Matthews et al., 2011; Peluso et al., 2009; Ruhé et al., 2012), while several others found decreased activation (Fales et al., 2008; Moses-Kolko et al., 2010). Furthermore, only a single study found an association of depression with the contrast of amygdala activation between fearful-face and to neutral-face conditions (Fales et al., 2008). Multiple other studies did not find any significant differences (Supplementary Section S1). With regard to the prefrontal cortex, reports have also been inconsistent. Some found decreased activations in the DLPFC, dorsomedial prefrontal cortex (DMPFC), orbitofrontal cortex or superior frontal gyri – primarily when emotional processing was implicit (Bürger et al., 2017; Fales et al., 2008; Kerestes et al., 2012; Matthews et al., 2011; Moses-Kolko et al., 2010; Ruhé et al., 2012; Wackerhagen et al., 2020). Several other studies, however, report increased activations of the DLPFC, DMPFC and in frontal gyri (Luo et al., 2018; Norbury et al., 2010; Powers et al., 2017). Samples in most previous studies had *N* < 50 cases, which may be the reason for inconsistent results.

Beyond studies of activation, some evidence indicates depression-related disrupted connectivity between the amygdala and the prefrontal cortex in response to fearful faces. For example, Moses-Kolko et al. (2010) applied Granger causality analysis and found that whereas top-down effective connectivity from left DMPFC to the left amygdala was present in controls, it was absent in depression. Kong et al. (2013) reported decreased functional connectivity between the amygdala and left rostral PFC, assessed with simple correlation analyses. Finally, Wackerhagen et al. (2020) applied the generalised psychophysiological interaction (gPPI) framework and revealed depression-related decrease in functional connectivity between the amygdala and right middle frontal gyrus. Numbers of depressed participants in these studies were still relatively small, respectively *N* = 28, *N* = 14 and *N* = 48.

In the current study we aimed to directly address the inconsistent results in the past literature (see above and Supplementary Section S1) by analysing brain activations and effective connectivity between the amygdala and the DLPFC in response to fearful faces in the large brain imaging subsample of the deeply-phenotyped Generation Scotland cohort (Habota et al., 2021). We hypothesised that, compared to controls, participants with lifetime (including current) experience of depression would be characterised by increased activation of the amygdala and by altered (increased or decreased, given mixed past evidence) activation of the DLPFC, when viewing fearful faces. Because the amygdala receives processed information from the visual areas, we also hypothesised that higher amygdala activity could be driven in part by stronger effective connectivity from these areas (Adolphs and Spezio, 2006; Pessoa and Adolphs, 2010). Finally, we aimed to address the outstanding question regarding the limbic-prefrontal connectivity in depression and define whether altered brain activations may be driven by changes in ‘bottom-up’ (amygdala to DLPFC) or ‘top-down’ (DLPFC to amygdala) effective connections (Elliott et al., 2011; Groenewold et al., 2013; Rive et al., 2013).

## Materials and Methods

### Participant Sample

In total, brain imaging and diagnostic data were available for *N* = 1,058 participants from the Generation Scotland cohort (*N* = 544 from Aberdeen, *N* = 514 from Dundee) (Habota et al., 2021). Of these, *N* = 47 had a diagnosis of current depression, *N* = 270 had a diagnosis of past depression, and *N* = 741 were classed as controls. Participants were classed as having had experience of depression within their lifetime (LMDD) if they met criteria for either current or past depression. Diagnoses were established with the research version of the Structured Clinical Interview for DSM Disorders during a face-to-face assessment (First et al., 2002), and were based on DSM-IV (American Psychiatric Association, 2000). Each participant also completed the Quick Inventory of Depressive Symptomatology (QIDS) scale (Rush et al., 2003) to obtain a measure of depressive symptom severity over seven days immediately prior to assessment. The study received ethical approval from the NHS Tayside committee on research ethics (reference 14/SS/0039). All participants gave written informed consent.

### Brain Imaging

#### Scanning Details

Structural and functional brain imaging was performed with a Philips Achieva 3T scanner in Aberdeen, and with a Siemens Prisma-FIT 3T scanner in Dundee. Scanning parameters were generally similar between the two sites. Please see Supplementary Section S2.1 for further details.

#### Task Details

The in-scanner task targeted implicit emotional processing. Participants were presented with blocks of trials with images of either neutral or fearful emotional faces from the NimStim dataset (Tottenham et al., 2009), and were required to identify the gender of each face (male or female). The recorded behavioural measures included reaction times and response correctness (Supplementary Section S2.2 provides further details).

### Preprocessing and Quality Control

Preprocessing was completed using default settings in SPM12 (https://www.fil.ion.ucl.ac.uk/spm/software/spm12). Supplementary Section S2.3.1 provides complete details of the preprocessing steps and parameters.

Quality control was performed in three main steps: 1) Detection of artefactual volumes with ArtRepair toolkit (Mazaika et al., 2009) and exclusion of participants with fractions of artefact volumes above threshold; 2) Visual inspection of representative functional and structural volume slices and exclusion of participants with anatomical abnormalities, normalisation or signal drop-out problems, and 3) Exclusion of participants with low task performance accuracies in the scanner. Supplementary Section S2.3.2 provides further details of quality control.

### Behavioural Data Analysis

Behavioural measures for each participant included reaction times and accuracies (fractions of correct responses) for the entire task, and separately for neutral-face and fearful-face conditions; as well as differences in mean reaction times between fearful-face and neutral-face conditions; and fractions of trials with missed responses. To test associations with depression, linear regression models were fit for each behavioural measure with either LMDD status or QIDS score entered as the main predictor variable; age, sex and site as nuisance covariates; and the tested behavioural measure as the response variable (function *fitlm* in MATLAB R2018a, MathWorks Inc).

### Brain Activation Analyses

#### First-level Brain Activation Analysis

Onsets and durations of blocks with neutral and fearful face stimuli were entered as regressors for the two task conditions in the first-level general linear model (GLM) design matrix. Six within-scanner movement parameters (three translation and three rotation parameters) were entered as nuisance covariates. Serial correlations were accounted for with a first-order autoregressive model. The data were high-pass filtered with a 128-second cut-off. As in Rupprechter et al., (2020), first-level relative masking threshold was set to 0.4 to increase the brain area included in the second-level analysis, and an explicit SPM mask for intracranial volume (ICV) was applied to limit the analyses to voxels within the brain.

#### Group-level Whole-brain Activation Analyses

Contrast images from the first-level analysis (Neutral > Baseline, Fearful > Baseline, Neutral > Fearful, Fearful > Neutral) were entered into the second-level (group) analyses, aiming to investigate associations with LMDD in any contrast. Age, sex and site (Aberdeen or Dundee) were entered as nuisance covariates in all second-level analyses. For exploratory whole-brain analyses, the cluster-level family-wise error (FWE) corrected threshold was specified as *P* < 0.05, with the whole-brain (cluster-forming) threshold set to uncorrected *P* < 0.0001. To investigate whether any differences associated with LMDD are related to severity of current depression symptoms, additional analyses with QIDS score as the main predictor variable (instead of LMDD) were performed 1) in the entire sample, 2) in the sample of LMDD participants, and 3) in the sample of participants with current major depressive episode (Table 1). To additionally investigate whether antidepressant medication may have influenced the findings, all analyses with significant results were rerun with an added binary covariate indicating antidepressant medication status.

**Table 1.** Summary demographic characteristics of the sample included in the analyses of behavioural measures and brain activations.

| Characteristic | Controls | LMDD |
| --- | --- | --- |
| Size (*N*) | 664 | 290 |
| Current MDE (*N*) | – | 41 |
| Sex (male / female) | 302 / 362 | 73 / 217 |
| Age (years) | 59.96 (9.98) | 56.26 (10.25) |
| QIDS (score) | 3.51 (2.14) | 6.97 (4.85) |
| Medicated (*N*) | 24 (3.6%) | 108 (37.2%) |

**Note:** MDE – major depressive episode. Participants were considered medicated if they had an antidepressant prescription at the time of the scan. Standard deviations are in parentheses.

#### Group-level ROI Activation Analyses

Same group-level analyses as described above for whole-brain activations were also performed for two predefined ROIs with small volume correction (SVC). The two ROIs were the bilateral amygdala and the bilateral DLPFC. ROI masks were defined according to the Talairach Daemon atlas within the Wake Forest University School of Medicine PickAtlas tool (WFU PickAtlas, Maldjian et al., 2003; Lancaster et al., 2007). DLPFC ROI was specified as a combination of bilateral Brodmann areas 9 and 46. Please see Supplementary Section S2.5 for further details and rationale for the definition of the two ROIs. Despite the lowered first-level masking threshold (from SPM default 0.8 to 0.4), signal was absent in a small caudal area of the amygdala ROI for some participants, and this area was not included in group-level analyses. Supplementary Figures S1 and S2 illustrate the amygdala ROI and the signal dropout area.

Apart from the above SVC analyses, we also investigated associations of clinical variables with mean bilateral amygdala activations at neutral- or fearful-face task conditions. Mean amygdala activations were estimated with MarsBaR toolkit (Brett et al., 2002). Statistical analyses were performed by fitting linear regression models with age, sex and scan site entered as nuisance covariates. As above, all analyses with significant results were rerun with an additional covariate indicating antidepressant medication status.

### Effective Connectivity Analyses

#### Effective Connectivity Analysis Overview

Dynamic causal modelling (DCM, Friston et al., 2003; Stephan et al., 2010) was applied to investigate associations of LMDD with effective connectivity between the visual cortex (V1) and the amygdala, as well as connectivity between the amygdala and the DLPFC. A bilinear DCM with three regions (left V1, left amygdala, left DLPFC), one state per region, no stochastic effects and mean-centred inputs was specified. A fully-connected model with nine connections (including inhibitory self-connections at each region) was assumed. Fearful and neutral trial blocks served as driving inputs to the visual cortex and could also modulate any of the nine endogenous connections (‘B’ matrix specified as ones for the two conditions). There were 27 free model parameters including nine connections and 18 modulatory inputs. Connections from V1 to amygdala and from amygdala to DLPFC were assumed to be direct (Adolphs and Spezio, 2006; Folloni et al., 2019; Marek et al., 2013; Pessoa and Adolphs, 2010; Ray and Zald, 2012), while connections between V1 and the DLPFC were assumed to be largely indirect (Paneri and Gregoriou, 2017). Figure 1 illustrates the DCM model structure. The DCM regions were defined according to the peak differences between LMDD and control participants in the SVC analyses and were in the left hemisphere (please see the results section below). Supplementary Section S2.6 provides further rationale for the DCM model specification, as well as details of the DCM ROI definition and the ROI time-series extraction.

**Figure 1.**
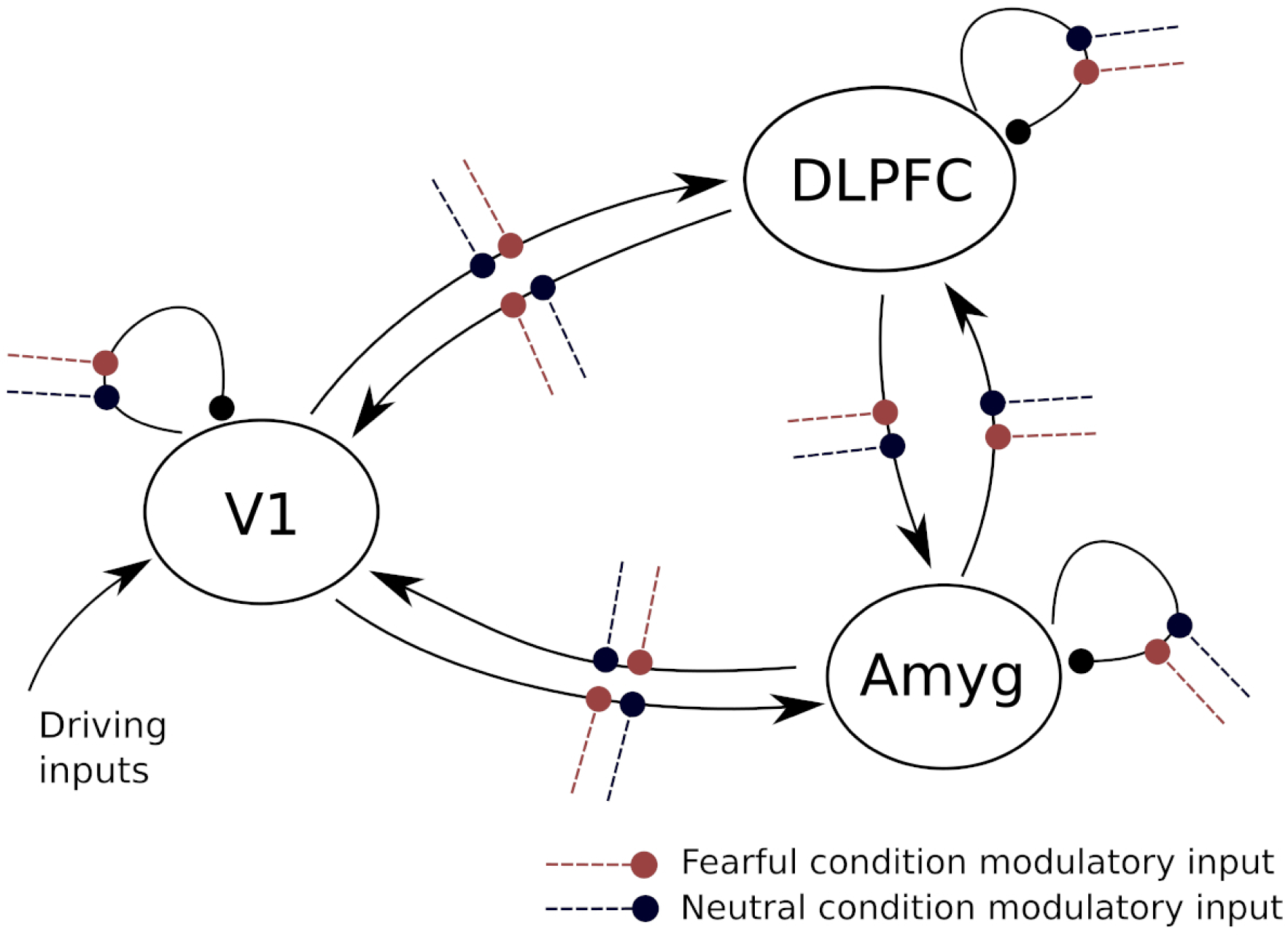
DCM model structure. Effective connectivity model structure specified for first-level fitting.

#### First-level Effective Connectivity Model Fitting

The full DCM model (Figure 1) was fitted for each participant using the standard Variational Laplace methodology implemented in SPM12, and percentage variance explained was estimated. Participants with at least 10% of time-series data variance explained by the model were included in the further group-level analyses, as in the previous work in our lab (Rupprechter et al., 2020; Zeidman, 2019).

#### Group-level Effective Connectivity Analyses

The parametric empirical Bayes (PEB) framework was used for group-level analyses of effective connectivity (Friston et al., 2016). Group-level PEB design matrix included a column of ones to model mean connectivity across the participants, and four zero-mean centred covariates – LMDD status, age, sex and scan site. PEB model was inverted to estimate model parameters and the model evidence (approximated by the free energy property). Bayesian model reduction was used to iteratively estimate different reduced PEB models with certain model parameters disabled, within an automatic greedy search procedure. Reduced models that were identified at the final iteration of the search procedure were combined using Bayesian model averaging (Friston et al., 2016; Penny et al., 2007). It was hypothesised that LMDD would be related to increased modulation of connection V1 to Amygdala, and altered modulation of connections Amygdala to DLPFC or DLPFC to Amygdala by the fearful-face task condition (above 0.5 posterior probability of relevant DCM parameters being non-zero). To check if any differences associated with LMDD are also related to the severity of current depression symptoms, an additional analysis with QIDS score as the main predictor variable (instead of the LMDD status) was performed. All effective connectivity analyses of LMDD were also rerun with an added binary zero-centred covariate indicating antidepressant medication status (similarly to group-level activation analyses).

## Results

### Participant Sample

*N* = 954 participants met the criteria to be included as either controls (*N* = 664) or LMDD cases (*N* = 290), and passed quality control. Summary demographic characteristics of the sample included in the group-level analyses are presented in Table 1. Supplementary Section S3.1 provides details of participant exclusions during quality control.

### Behavioural Results within Scanner

No significant associations were found between LMDD and either reaction times (complete task or two task conditions separately), accuracies, or fractions of missed trials (all *P* > 0.25). See Supplementary Section S3.2 for further details and for the results of analyses with QIDS scores.

### Brain Activation Results

#### Group-level Brain Activations

Brain activations in response to both neutral and fearful face stimuli in the entire sample covered multiple areas including bilateral occipital, fusiform, paracentral, precentral, prefrontal and right parietal cortices. SVC analyses revealed activations in the right amygdala and in bilateral DLPFC. The activations were generally consistent with those reported in the previous studies (Fusar-Poli et al., 2009). Please see Supplementary Section S3.3.1 for further details.

#### Differences in Whole-brain Activations in LMDD

For Neutral > Baseline contrast, exploratory whole-brain analyses revealed higher activations in LMDD (compared to controls) in left dorsal anterior cingulate cortex (ACC), bilateral precentral, right temporal and left parietal / occipital cortices. For Fearful > Baseline, higher LMDD-related activations were found in left DLPFC / precentral cortex, left primary somatosensory, right precentral and mid-cingulate cortices. For the details and illustration of the identified activation differences please see Table 3 and Figure 2A. No significant LMDD-related differences were found for Neutral > Fearful and Fearful > Neutral contrasts.

**Figure 2.**
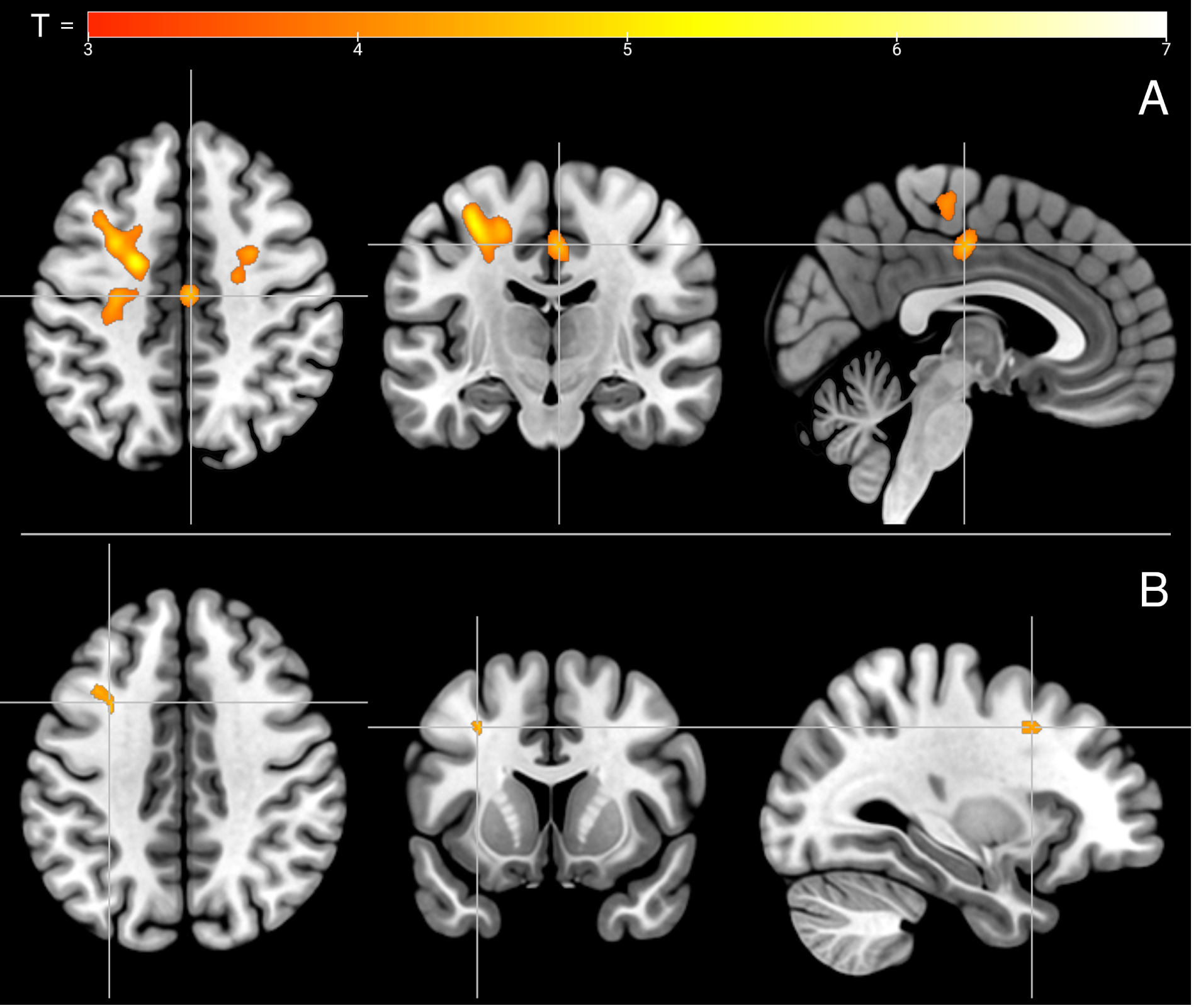
Increased brain activations in LMDD. Stronger activations in LMDD compared to controls for Fearful > Baseline contrast: (A) whole-brain with cluster-level FWE correction (*P* < 0.0001 uncorrected cluster-forming threshold, *P* < 0.05 cluster-level threshold), crosshair centred on mid-cingulate cortex (MNI 3 −16 44); (B) DLPFC SVC (*P* < 0.05 small-volume FWE correction), crosshair MNI coordinates −31 12 40.

**Table 2.** Summary demographic characteristics of the sample included in the analyses of effective connectivity.

| Characteristic | Controls | LMDD |
| --- | --- | --- |
| Size (*N*) | 474 | 214 |
| Current MDE (*N*) | – | 32 |
| Sex (male / female) | 230 / 244 | 53 / 161 |
| Age (years) | 60.09 (9.90) | 56.80 (10.14) |
| QIDS (score) | 3.45 (2.12) | 6.90 (4.84) |
| Medicated (*N*) | 15 (3.2%) | 85 (39.7%) |

**Note:** MDE – major depressive episode. Participants were considered medicated if they had an antidepressant prescription at the time of the scan. Standard deviations are in parentheses.

**Table 3.** Higher activations in LMDD compared to controls with whole-brain cluster-level correction and with DLPFC small volume correction.

| **Correction** | **Contrast** | **Region** | **Peak MNI** | **Cluster-level significance** | **Cluster extent** |
| --- | --- | --- | --- | --- | --- |
| *Whole-brain cluster-level* | Neutral<br>> Baseline | Left dorsal ACC | -18 30 30 | $P_{\text{FWE}} = 0.018$ | $k_E = 83$ |
| | | Left precentral | -36 18 40 | $P_{\text{FWE}} < 0.001$ | $k_E = 288$ |
| | | Right precentral | 26 2 42 | $P_{\text{FWE}} = 0.002$ | $k_E = 150$ |
| | | Right temporal | 46 -16 34 | $P_{\text{FWE}} = 0.001$ | $k_E = 192$ |
| | | Left parietal or occipital | -46 -62 22 | $P_{\text{FWE}} = 0.002$ | $k_E = 244$ |
| | Fearful<br>> Baseline | Left DLPFC or precentral | -32 -18 38<br>-28 6 44 | $P_{\text{FWE}} < 0.001$ | $k_E = 1020$ |
| | | Left primary somatosensory | -6 -30 56 | $P_{\text{FWE}} < 0.001$ | $k_E = 225$ |
| | | Right precentral | 22 -8 42 | $P_{\text{FWE}} = 0.002$ | $k_E = 149$ |
| | | Mid-cingulate | 2 -16 44 | $P_{\text{FWE}} = 0.016$ | $k_E = 87$ |
| *Amygdala small-volume* | Fearful<br>> Baseline | Left amygdala | -22 -10 -10 | $P_{\text{FWE}} = 0.031$ | $k_E = 4$ |
| | Fearful<br>> Neutral | Left amygdala | -24 -8 -10 | $P_{\text{FWE}} = 0.038$ | $k_E = 2$ |
| *DLPFC small-volume* | Neutral<br>> Baseline | Left DLPFC | -36 18 40 | $P_{\text{FWE}} = 0.001$ | $k_E = 44$ |
| | | | -18 30 30 | $P_{\text{FWE}} = 0.003$ | $k_E = 31$ |
| | Fearful<br>> Baseline | Left DLPFC | -30 8 42 | $P_{\text{FWE}} = 0.002$ | $k_E = 33$ |

Clusters of higher activation in right precentral and mid-cingulate cortices in LMDD for Fearful > Baseline contrast (Table 3) were no longer significant when antidepressant medication status was added as a covariate, but differences in other areas remained. The LMDD-related differences were not found in the analyses of QIDS scores, either in the entire sample, in the sample of LMDD participants only, or in the sample of participants with current MDD. This indicates that the differences associated with LMDD are not related to the severity of current depression symptoms. Please see Supplementary Sections S3.3.2-S3.3.3 for additional details.

#### Differences in Amygdala ROI Activation in LMDD

Analyses with the amygdala SVC revealed stronger activation in LMDD, compared to controls, in a small cluster in the left amygdala for Fearful > Baseline (*k _E_* = 4) and Fearful Neutral (*k _E_* = 2), but not for other contrasts (Table 3). These results were no longer significant when antidepressant medication status was added as a covariate.

Analysis with MarsBaR toolkit revealed that there was a higher mean bilateral amygdala activation in LMDD, compared to controls, in the fearful-face condition (*β* = 0.0715 *, P* = 0.0314). LMDD association with mean activation in the neutral-face condition was in the same direction but did not reach significance (*β* = 0.0601 *, P* = 0.0727). There was no significant association of LMDD with the contrast of amygdala activation between the fearful-face and neutral-face conditions. Figure 3 illustrates the amygdala activation compared between LMDD cases and controls. LMDD-related difference in the fearful-face condition remained significant when additionally correcting for antidepressant medication (*β* = 0.0725 *, P* = 0.0478).

**Figure 3.**
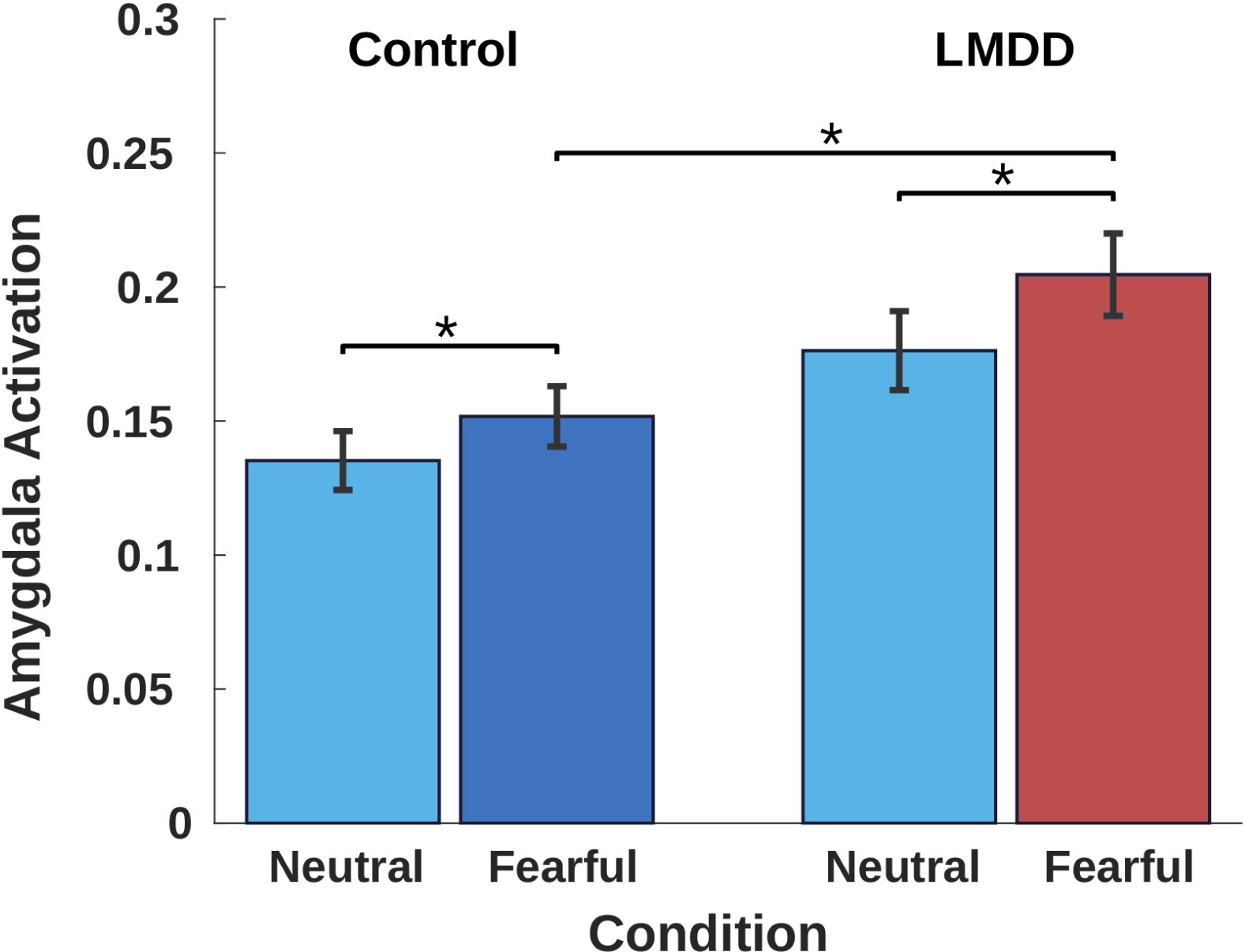
Amygdala activation levels. Mean bilateral amygdala activations (beta values estimated in the first-level analysis) in controls and LMDD cases for neutral-face and fearful-face conditions. Error bars represent standard errors of the mean. Stars indicate significant differences at *P* < 0.05. Significance calculated with paired *t*-tests (between conditions) and with linear regression (between groups, corrected for age, sex and site). Control and LMDD groups only significantly differed in levels of amygdala activation at fearful-face condition (*β* = 0.0715 *, P* = 0.0314).

No significant associations of amygdala activation with QIDS scores were found in the analyses with amygdala SVC, or in analyses of MarsBaR-derived mean amygdala activations, either in the entire sample or separately in the LMDD participants (all *P* > 0.21). When control participants were analysed separately, results revealed negative correlations of QIDS scores with mean bilateral amygdala activations in both fearful-face and neutral-face conditions (respectively *β* = −0.0898 *, P* = 0.0193 and *β* = −0.0819 *, P* = 0.0337). Please see Supplementary Section S3.3.3 for additional details.

#### Differences in DLPFC ROI Activations in LMDD

Analyses with DLPFC SVC revealed small clusters of increased activation in LMDD (compared to controls) in left DLPFC for Neutral > Baseline (*k _E_* = 44 and *k _E_* = 31) and Fearful > Baseline contrasts (*k _E_* = 33) (Table 3, Figure 2B). These differences were not found in the analyses of symptom severity (QIDS scores). Please see Supplementary Sections S3.3.2-S3.3.3 for further details.

### Effective Connectivity Results

#### Participant Sample in Effective Connectivity Analyses

Of *N* = 954 participants in total, *N* = 185 were excluded because they did not have sufficient activation in at least one of the three ROIs specified in the DCM (Supplementary Section S2.6). Another *N* = 81 participants were excluded due to having less than 10% of the variance of time-series data explained by the fitted DCM model (Rupprechter et al., 2020; Zeidman, 2019). Demographic characteristics of the sample in effective connectivity analyses are presented in Table 2. Compared to those included, participants who were excluded from the DCM analyses on average had significantly higher fractions of artefactual volumes (1.42% compared to 1.08%, *t* (445) = 2.32 *, P* = 0.0206), but there were no differences in terms of any demographic or clinical characteristics, or any measures of task performance within the scanner (Supplementary Section S3.4.1 and Table S2 provide further details).

#### Group-level Effective Connectivity

At the group-level, all endogenous (task-independent) connections were found to have high probability. Connections from V1 were found to be excitatory, while all other connections (from amygdala or the DLPFC) were found to be inhibitory. The neutral-face task condition positively modulated (increased strength) the V1 incoming connections and the Amygdala to DLPFC connection, and negatively modulated (decreased strength) the V1 to DLPFC connection. The fearful-face task condition negatively modulated the V1 inhibitory self-connection and the V1 to DLPFC connection, and positively modulated the amygdala and DLPFC self-connections and the V1 incoming connections. Figure 4 illustrates the DCM model after fitting. See Supplementary Tables S15-S16 for specific estimated connectivity model parameters.

**Figure 4.**
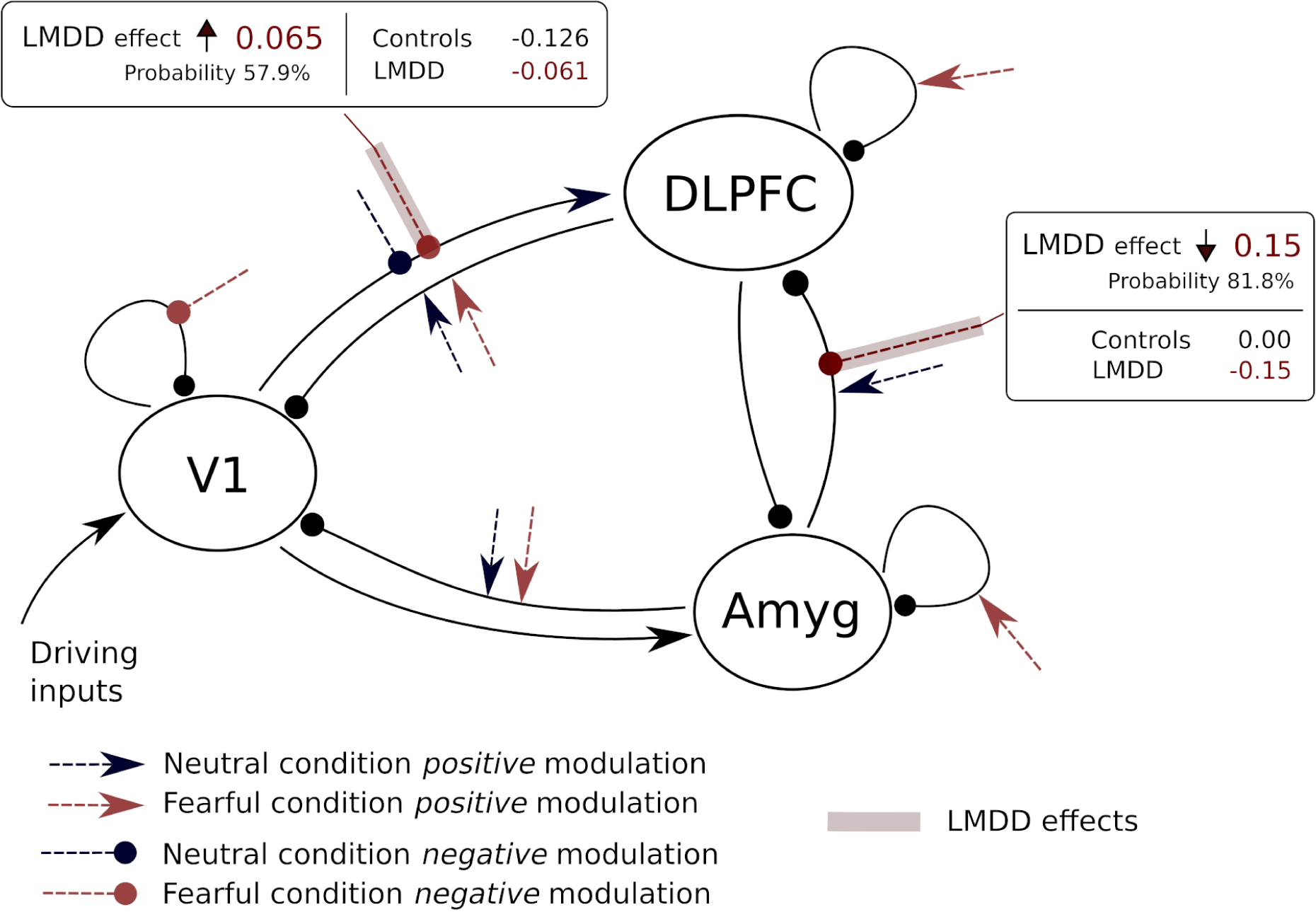
Fitted DCM model with the estimated changes related to LMDD. Only connections and modulatory inputs estimated to be plausible are shown. Excitatory connections are denoted with arrows, inhibitory connections are denoted with circles. Boxes describe changes in LMDD.

#### Changes in Effective Connectivity in LMDD

LMDD was found to be related to negative modulation of the Amygdala to DLPFC connection (effect −0.15, probability 0.82), and to reduced negative modulation of the V1 to DLPFC connection (effect 0.065, probability 0.58), both in the fearful-face task condition. This indicates stronger signalling from V1 to DLPFC, and stronger inhibition from the amygdala to the DLPFC, in response to fearful faces in LMDD compared to controls. No additional associations of LMDD were discovered when group-level PEB analyses were repeated with estimation of only endogenous connection parameters (‘A’ matrix, nine parameters), or only modulatory inputs (‘B’ matrix, 18 parameters) (Zeidman et al., 2019). Results did not change when antidepressant medication status was included as a covariate in the PEB design matrix of the full DCM model. Supplementary Section S3.4.2 provides further details.

When LMDD status was replaced with QIDS scores, no significant associations were found in the full DCM model, indicating that the changes in LMDD were not related to severity of current depression symptoms. Please see Supplementary Section S3.4.3 for further results with separate estimation of connectivity and modulation parameters.

## Discussion

### Summary of Results

Our results indicate that experience of depression within the lifetime is characterised by *increased* activations in the left DLPFC and bilateral precentral cortices in response to both fearful and neutral face stimuli, and also in the left amygdala in response to fearful faces. Moreover, as compared to controls, LMDD is associated with stronger amygdala to DLPFC *inhibitory* connectivity when viewing fearful faces. These findings were not attributable to either severity of acute depressive illness or to antidepressant medication status, and make a significant contribution to addressing ambiguities in the previous literature.

### Hyperactivation to Fearful Faces in LMDD

Results of previous studies were inconsistent and only some have found significant differences in amygdala activation in response to fearful faces in depression (Supplementary Section S1 and Table S1). With *N* = 290 cases and *N* = 664 controls, our sample was substantially larger than those of the previous studies, where typically *N* < 50 depressed or remitted-depressed participants were investigated. It is possible that inconsistency in the previous findings may have been due to the small sample sizes, insufficient to detect low-magnitude effects such as reported here. Because our sample combined cases of both current and past depression, the results indicate that higher amygdala reactivity may be a marker of either vulnerability or the lasting effects of the experience of depression, rather than of the current depressive state. This is corroborated by the previous studies which found higher fearful-face-related amygdala activation in participants at high risk for depression due to either parental mental health history (Monk et al., 2008), high neuroticism (Chan et al., 2009), vulnerability to depression-related cognition (Zhong et al., 2011), or past depression (Korgaonkar et al., 2019). Hyperactive amygdala has been suggested as one of the core mechanisms of negative bias in several prominent neurocognitive theories of MDD (DeRubeis et al., 2008; Disner et al., 2011; Mayberg, 1997; Roiser and Sahakian, 2013), due to its fundamental role in emotional processing (Phelps, 2006; Phelps and LeDoux, 2005; Sergerie et al., 2008). Our results lend credence to these theories, specifically in the context of processing emotional faces.

With regard to the DLPFC, Norbury et al. (2010) previously found increased fearful-face-related activation in remitted depression (compared to healthy controls). Powers et al. (2017) and Luo et al. (2018) found positive correlations of depression symptom severity (BDI scores) with activations of the DMPFC or frontal gyri. Studies by Ruhé et al. (2012), Matthews et al. (2011), and Fales et al. (2008), however, found decreased fearful-face-related DLPFC activations in current MDD. These contrasting results could be due to differences in diagnostic criteria: whereas more severe current MDD could lead to decreased DLPFC activation, lifetime experience (past depression) could potentially lead instead to compensatory hyperactivation – as seen in our study. Secondary evidence which indirectly supports this possibility is that transcranial stimulation of the DLPFC can be an effective treatment (Razza et al., 2020; Teng et al., 2017). DLPFC is overall a large region and it is also possible that changes in its activation are heterogeneous – with hyperactivation in some sub-regions but deactivation in others.

Despite the relatively large sample size, we only found significant depression-related differences in the amygdala and the DLPFC when fearful face processing was compared with baseline, but not when compared with neutral face processing (with an exception for a small left amygdala cluster in SVC analyses). Differences in the fearful to neutral face contrast in depression were only found in a single past study in the amygdala (Fales et al., 2008), and in three studies in the DLPFC (Fales et al., 2008; Kerestes et al., 2012; Powers et al., 2017). The depressive negative bias likely affects processing of both emotional and neutral faces – with a tendency for neutral faces to be interpreted as negative, which can in part explain the lack of depression-related differences in the contrast of fearful to neutral faces (Bourke et al., 2010; Elliott et al., 2011; Stuhrmann et al., 2011). It is possible that in depression the amygdala is hyperactive to all faces, simply because any face can take negative expressions, but then is additionally hyperactive in proportion to the intensity of negative expression. Future studies could test this by using carefully prepared images of facial expressions with varying degrees of intensity. Amygdala hyperactivity could also depend on the severity of current symptoms, or presence of co-morbid anxiety.

Apart from increased activations in the amygdala and the DLPFC, we also found increased fearful-face-related mid-cingulate activation in LMDD, although this result did not survive correction for antidepressant medication status. One possibility is that antidepressant intake may have driven the increased mid-cingulate activation. This would be consistent with the results of studies by Ruhé et al. (2012) and Bürger et al. (2017), who found that current MDD was related to decreased rather than increased cingulate activation in response to fearful faces. Conversely, it is possible that correction for antidepressant medication may have simultaneously adjusted for the effects of more severe depression (medicated participants are likely to be more depressed), potentially including the increased mid-cingulate activation. This could be consistent with the previous findings of positive correlations of cingulate activation with self-reported depression symptom severity (Bürger et al., 2017; Luo et al., 2018). From a functional perspective, increased cingulate activity found in our study could represent higher cognitive efforts deployed to stay focused on the task in LMDD, which may in turn have helped maintain correct task performance in the presence of fear-related stimuli (Cavanagh and Shackman, 2015; Shackman et al., 2011). Future studies are necessary to clarify whether increased cingulate activity may be driven by depression modulated by antidepressant medication, and what the exact role of this hyperactivity is with regard to behavioural performance.

### Effective Connectivity of the Amygdala and DLPFC in LMDD

Our analyses of effective connectivity revealed that the amygdala exerted inhibitory influence over the DLPFC, and that this inhibitory influence was increased in LMDD (compared to controls), specifically in response to fearful face stimuli (Figure 4). These results are generally consistent with the studies of Kong et al. (2013) and Wackerhagen et al. (2020). Moses-Kolko et al. (2010), however, found altered ‘top-down’ (left DMPFC to left amygdala), but not ‘bottom-up’ connectivity when processing fearful faces – somewhat contrary to our results. We note that these discrepant findings could be due to differences in diagnostic inclusion criteria: whereas we investigated relatively older adults of both sexes, Moses-Kolko et al. focused on younger individuals with postpartum depression.

Prominent neurocognitive theories of depression suggest imbalance in activities between prefrontal and limbic areas (DeRubeis et al., 2008; Disner et al., 2011; Mayberg, 1997), however the origin of this imbalance has not yet been clearly defined. According to one theoretical view, depressive symptomatology is primarily a result of lower-level biases in information processing (perception and reinforcement learning), subserved mainly by hyperactive limbic structures including the amygdala (Roiser et al., 2012; Roiser and Sahakian, 2013). In that view, deficits in cognitive control represent a secondary vulnerability. An alternative suggestion, however, is that the origin of negative biases is in inability to effectively regulate and inhibit processing of negative emotional information, which is mainly related to deficits in cognitive control and hypoactive prefrontal cortical areas (Gotlib and Joormann, 2010; Joormann, 2010). In this study we only found evidence for increased inhibitory influence from the amygdala to the DLPFC, but no altered connectivity from the DLPFC to the amygdala. This provides support for the former theory – that depressive symptoms are mainly a result of ‘bottom-up’ rather than ‘top-down’ biases in emotional processing, and that these biases may be subserved by an inhibitory, rather than an excitatory, attention-orienting modulation of the DLPFC by the amygdala (Stolicyn et al., 2017). This finding implies that at least some subtypes of depression may be susceptible to treatments which target lower-level emotional information processing biases – for example reinforcement-based attention training (Jonassen et al., 2019). Future work should investigate whether limbic-prefrontal connectivity deficits could represent a prognostic marker of response to such interventions that target lower-level perceptual and attentional biases.

It should be noted that the results of our effective connectivity analyses do not immediately explain the observed changes in activation patterns in LMDD. Specifically, it remains an open question how increased inhibitory input from the amygdala may translate to increased activation of the DLPFC. One example explanation which could reconcile these findings is that the DLPFC activation may fluctuate throughout the different task trial stages. The DLPFC may, for example, be inhibited by the amygdala at stimulus onset, and thus be less active specifically at this trial stage, but may then accumulate stronger inputs from other areas such as V1 and become hyperactive by the time of response execution. Activity fluctuations may average out over the course of entire trial blocks, resulting in an apparent DLPFC hyperactivity as observed here. We did not investigate this possibility here due to the block design and the relatively low numbers of trials per condition – however it remains an open avenue for future studies with event-related task designs. An alternative explanation could be that the DLPFC may be receiving excitatory inputs from brain areas outside our considered DCM model, which outweigh the inhibition received from the amygdala. One such area, for example, could be the ACC, due to its connectivity with the DLPFC (Bubb et al., 2018). To investigate this possibility, future large-sample studies could consider more complex connectivity models with higher numbers of regions that could potentially modulate activation patterns of the DLPFC.

### Limitations and Conclusion

A distinct advantage of our study is the large sample size, however several limitations should be mentioned. First of all, the identified depression-related changes were small and no differences in behavioural measures indicative of a negative bias were observed. A previous study with out-of-scanner cognitive tasks in the imaging subsample of the Generation Scotland cohort (*N* = 1109, including most participants from the current study) did find evidence of a subtle negative bias effect associated with depression symptoms (de Nooij et al., 2022). It is possible that the within-scanner gender identification task studied here was sufficient to induce negative bias-related effects at the neural but not at the behavioural level, in contrast to the out-of-scanner tasks studied in de Nooij et al. (2022). With regard to the small effect sizes, they are generally characteristic of larger-sample studies (Button et al., 2013), however in our study this could also be because most case participants had past rather than current depression, and because average severity of symptoms was relatively low. Moreover, a substantial proportion of participants were taking antidepressant medication (Tables 1-2). We thus cannot claim that the results extend directly to unmedicated patients with more severe symptoms. More severely depressed patients are more likely to be medicated, and hence it is generally difficult to disentangle effects of medication and depression severity. Further to that, our dataset did not involve longitudinal assessment of symptoms, and we did not test altered brain activations or connectivity as prognostic biomarkers. Future studies should assess larger, longitudinal samples of participants with current and more severe clinical depression, ideally of younger age, in order to identify changes which may be relevant as diagnostic or prognostic biomarkers (Godlewska et al., 2016; Williams et al., 2015).

In summary, the current study indicates that lifetime experience of depression is associated with increased activation of the amygdala and with increased inhibitory influence of the amygdala over the DLPFC in response to fearful faces. This helps reduce ambiguity in the previous literature and provides support to the neurocognitive theories which state that the depressive negative bias may be underpinned by changes in ‘bottom-up’ connectivity from limbic to higher cortical areas.

## Data Statement

Data from the Generation Scotland dataset are available through application to the Generation Scotland Access Committee and Edinburgh DataVault (https://doi.org/10.7488/8f68f1ae-0329-4b73-b189-c7288ea844d7). For further information, please see Habota et al. (2021).

## Data Availability

Data from the Generation Scotland dataset are available through application to the Generation Scotland Access Committee and Edinburgh DataVault (https://doi.org/10.7488/8f68f1ae-0329-4b73-b189-c7288ea844d7).

## Acknowledgements

For further details regarding the NimStim dataset of facial expressions please see https://danlab.psychology.columbia.edu/content/nimstim-set-facial-expressions.

Development of the MacBrain Face Stimulus Set was overseen by Nim Tottenham and supported by the John D. and Catherine T. MacArthur Foundation Research Network on Early Experience and Brain Development. Please contact Nim Tottenham at for more information concerning the stimulus set. The current work has made use of the resources provided by the Edinburgh Compute and Data Facility (ECDF) (http://www.ecdf.ed.ac.uk/). We would like to thank all Generation Scotland research volunteers who took part in the current study for contributing their time and effort. We would also like o thank all research assistants, clinicians and technicians for their valuable contributions to collection of the data.

