## Supplementary Material for "Disrupted Limbic-Prefrontal Effective Connectivity in Response to Fearful Faces in Lifetime Depression"

Aleks Stolicyn[1], Mathew A. Harris[1], Laura de Nooij[12], Xueyi Shen[1], Jennifer A. MacFarlane[345], Archie Campbell[6], Christopher J. McNeil[57], Anca-Larisa Sandu[57], Alison D. Murray[57], Gordon D. Waiter[57], Stephen M. Lawrie[1], J. Douglas Steele[35], Andrew M. McIntosh[156], Liana Romaniuk*[15], Heather C. Whalley*[156]

***** These authors share joint senior authorship

1. Division of Psychiatry, Centre for Clinical Brain Sciences, University of Edinburgh, Kennedy Tower, Royal Edinburgh Hospital, Edinburgh EH10 5HF, United Kingdom
2. Donders Institute for Brain, Cognition and Behaviour, Radboud University Medical Center, Nijmegen 6525 EN, Netherlands
3. Division of Imaging Science and Technology, School of Medicine, University of Dundee, Dundee DD1 9SY, United Kingdom
4. Department of Medical Physics, NHS Tayside, Dundee DD2 1UB, United Kingdom
5. SINASPE Consortium, https://www.sinapse.ac.uk
6. Centre for Genomic and Experimental Medicine, Institute of Genetics and Cancer, University of Edinburgh, Edinburgh EH4 2XU, United Kingdom
7. Aberdeen Biomedical Imaging Centre, Institute of Medical Sciences, University of Aberdeen, Aberdeen AB25 2ZN, United Kingdom

Corresponding author:

Aleks Stolicyn
Division of Psychiatry, Centre for Clinical Brain Sciences, University of Edinburgh
Kennedy Tower, Royal Edinburgh Hospital, Morningside Park, Edinburgh EH10 5HF, UK.


#### S1. BACKGROUND

A brief literature survey was conducted to identify existing evidence of changes in brain activation in response to fearful face stimuli. PubMed was searched with terms '*depression fearful face fMRI*' and '*depression fearful face connectivity*' in late 2020. Studies were selected for further review if they met the following criteria:

1) An adult human participant sample was investigated;

2) Functional brain imaging (except for electroencephalography) was employed;

3) Stimuli of faces with fearful emotional expressions were used in the task paradigm applied within the scanner;

4) Associations of either current or past major depressive disorder (MDD) were investigated, either by comparing cases and controls, or by assessing correlations of depression symptom severity levels with brain activations.

Overall, 30 studies fitting the above criteria were identified. Please see Table S1 for summary details of the identified studies. Cells highlighted in light red in Table S1 outline differences in brain activation in response to fearful faces in depression. Depressed or remitted depressed participants were studied in 28 of the 30 studies. Another 2 studies investigated correlations of brain activations with Beck Depression Inventory (BDI) scores in non-clinical participants. Most studies (22 out of 30) assessed differences in whole-brain activations. Regions of interest which were most frequently investigated were the amygdala (20 of 30 studies) and the prefrontal cortex (six studies). Despite strong focus on the amygdala in the literature, depression-related differences in processing fearful faces (compared to either baseline or neutral faces) have only been found in seven studies.[1–7] Most have found increased activation,[1–6] although decreases were reported in two studies.[6,7] Ten studies identified effects of depression in the prefrontal cortex (mostly within whole-brain analyses). Seven have found decreased activations in the DLPFC, dorsomedial prefrontal cortex (DMPFC), orbitofrontal cortex (OFC) or superior frontal gyri – primarily where emotional processing was implicit (Table S1).[3,4,6–10] Three other studies, however, to the contrary found increased activations in the DLPFC, DMPFC and in frontal gyri.[11–13]

#### S2. MATERIALS AND METHODS

##### *S2.1 Brain Scanning Details*

Structural and functional brain imaging was completed with a Philips Achieva 3T scanner in Aberdeen, and with a Siemens Prisma-FIT 3T scanner in Dundee. In both Aberdeen and Dundee, volumes in functional brain scans had 32 axial slices, and were acquired with repetition time of 1560 ms, flip angle of 70 degrees, field of view of 217 mm, matrix size of 64 × 64, and voxel size of 3.4 × 3.4 × 4.5 $mm^3$. Echo time was 26 ms in Aberdeen and 22 ms in Dundee. For further details of both structural (T1-weighted) and functional magnetic resonance imaging sequences please see Habota et al. (2021).[14]

##### *S2.2 Behavioural Task Details*

Participants were presented with face images with either neutral or fearful expressions from the NimStim dataset[15] and were required to identify the gender of the face (male or female). There were three blocks of trials with neutral faces and three blocks with fearful faces, presented interchangeably. The first block always consisted of neutral faces. Each block had six trials (three with female faces and three with male faces, presented randomly), with 36 trials in total (18 with neutral faces and 18 with fearful faces). Each trial lasted 3.5 seconds with a 0.5 second inter-trial interval (24 seconds per block). There was an 8.5 second interval between consecutive blocks. Participant behavioural measures (reaction times and correct /

error responses) were recorded during the session.

#### *S2.3 Preprocessing and Quality Control*

##### S2.3.1 Preprocessing Steps

Default SPM12 (https://www.fil.ion.ucl.ac.uk/spm/software/spm12) settings were used in the preprocessing steps. The preprocessing steps were followings:

(1) Realignment of functional (echoplanar imaging, EPI) volume time-series to the time-series mean and reslicing for each participant;

(2) Co-registration of the realigned functional volumes with T1-weighted structural scans;

(3) Segmentation and bias-correction of participant T1-weighted scans;

(4) Normalisation of segmented and bias corrected T1-weighted scans to the standard MNI template through non-linear warping, recording of normalisation parameters;

(5) Normalisation of the co-registered functional (EPI) volumes with the transformation parameters recorded in step (4) above;

(6) Resampling of the functional (EPI) volumes at isotropic resolution of 2 mm and smoothing with a Gaussian kernel with full width to half maximum of 6 mm;

(7) Discarding of the first six volumes of the normalised functional (EPI) volume time-series for each participant.

##### S2.3.2 Quality Control Steps

Data quality control was completed with three primary steps:

(1) Detection of potentially artifactual echoplanar imaging (EPI) volumes and exclusion of participants with above-threshold fractions of artifact volumes (ArtRepair toolkit,[16] default recommended 0.5 mm scan-to-scan motion threshold, 10% artifactual

volumes threshold);

(2) Visual inspection of representative functional and structural volume slices and exclusion of participants with anatomical abnormalities, normalisation or substantial signal drop-out problems;

(3) Evaluation of participant behavioural performance at the task and exclusion of participants with low accuracy.

Visual inspection in step (2) above included checking 1) slices from the realigned mean functional volumes, 2) slices from the normalised structural volume overlaid over the representative normalised functional volume (to check functional co-registration and normalisation), 3) slices from the normalised structural volume overlaid over the MNI template (to check structural normalisation), and 4) slices from the first-level analysis mask (to check for signal drop-out).

In step (3) above, participants who missed (skipped) more than 10% of trials, or had an overall behavioural accuracy below 80% and above 20% were excluded. The threshold of 80% was chosen to strike a balance between excluding as few participants as possible, while at the same time ensuring that all included participants were focused on the gender identification task and paid attention to the stimuli. Participants with accuracy below 20% ($N = 15$) were kept following the assumption that they mixed the buttons necessary for correct responses.

##### *S2.4 Behavioural Data Analyses*

Differences in reaction times (RT) and gender identification accuracies between fearful-face and neutral-face conditions were assessed with paired sample *t*-tests, separately for cases and controls. Please see the methods section of the main text for details of assessment of group-level differences.

##### *S2.5 Brain Activation Analyses*

Bilateral amygdala and bilateral DLPFC region-of-interest (ROI) masks were defined according to the Talairach Daemon atlas within the Wake Forest University School of Medicine PickAtlas tool (WFU PickAtlas),[17] with corrected mapping from Talairach to MNI coordinates described in Lancaster et al. (2007).[18] DLPFC ROI was specified as a combination of bilateral Brodmann areas 9 and 46 with three-time repeated 3D dilation, as implemented in WFU PickAtlas, excluding out-of-brain areas. Brodmann areas 9 and 46 were specified because they are both involved in executive function and have been designated as the DLPFC ROI in past studies.[6,9,19,20] Each 3D dilation iteration expands the ROI by one 2-mm voxel in each direction and thus the DLPFC ROI was overall enlarged by 6 mm in each direction of each of the three axes. ROI dilation was applied because Brodmann areas generally represent thin cortical strips, and thus masks without dilation are too small in volume to capture the signal of interest.[17,21] Dilation also allows to take into account imperfect anatomical alignment between subjects after normalisation. Because amygdala represents an enclosed subcortical structure, its ROI was dilated only a single time.

##### *S2.6 Effective Connectivity Analyses*

###### S2.6.1 Effective Connectivity Analysis Overview

Dynamic causal modelling (DCM)[22,23] was applied to investigate associations of LMDD with effective connectivity between the visual cortex (V1) and the amygdala, and connectivity between the amygdala and the DLPFC. V1 was considered because the amygdala receives processed information from the visual areas,[24,25] and thus higher amygdala activation could be due to stronger signalling of fear-related information from these areas.[26] Connectivity between the amygdala and the DLPFC was modelled because the amygdala is known to have projections to prefrontal areas,[27–29] and indirect influence via neuromodulatory systems.[30–32] We considered that altered activations in these two regions could be driven at least in part by altered effective connectivity between them. Section S2.6.2 below describes definition of the regions of interest for the DCM model. Section S2.6.3 describes extraction of the ROI activation time-series for effective connectivity modelling.

###### S2.6.2 DCM Region of Interest Definition

For the amygdala and the DLPFC ROIs, activation search regions were first defined for each participant individually as combinations (logical ‘and’) of:

(1) Participant first-level analysis mask;

(2) Participant fearful-face activation significance map thresholded at $p < 0.10$, uncorrected (to exclude noisy voxels);

(3) Anatomical ROI mask (left amygdala or left DLPFC);

(4) 20-mm sphere centred at MNI [-22 -10 -10] for the amygdala or MNI [-36 16 38] for the DLPFC.

*Individual peak coordinates* were defined for each participant as coordinates of the most significant voxels in the activation search regions described above in the fearful-face condition (separately for the amygdala and DLPFC regions). For each participant, ROIs for time-series extraction were then defined as combinations (logical ‘and’) of items (1-3) above and 12-mm spheres centred at the identified participant *individual peak coordinates*. Definition of the participant *individual peak coordinates* enabled extraction of ROI time-series for a higher number of participants because the ROIs were tailored to maximise signal for each individual participant within the wider search regions. MNI coordinates in item (4) above were defined according to peaks of LMDD case-control differences in the fearful-face condition identified in SVC analyses (please see the results section of the main text). Please note item (4) above was only used to define the activation search regions, but not applied directly in the combinations to define the final ROIs. Spherical ROI size (12 mm) was defined to be consistent with the previous DCM work in our lab, which also assessed connectivity between cortical and subcortical structures.[33]

For the visual cortex, ROI was defined for each participant as a combination (logical 'and') of:

(1) Participant first-level analysis mask;

(2) Fearful-face activation significance map thresholded at $P < 0.10$, uncorrected (to exclude noisy voxels);

(3) 12-mm sphere centred at MNI [-18 -92 8].

The MNI coordinates in item (3) above were defined as coordinates close to the global peak activation in the fearful-face condition in the entire sample (which was in the left visual cortex), adjusted to be more anterior from the edge of the brain.

Participants who did not have any voxels active in response to fearful face stimuli in either of the three ROIs at significance level of $P < 0.10$ (uncorrected) did not have complete ROI time-series and were thus excluded from the DCM analyses.

##### S2.6.3 DCM Region Activation Time-series Extraction

First principal components of the pre-whitened, high-pass filtered and confounder-corrected (for six within-scanner movement parameters and serial correlations) time-series of the 12-mm ROI spheres defined above were extracted for each participant, as implemented in SPM12.

### S3. RESULTS

#### *S3.1 Quality Control Results*

Overall, $N = 86$ participants were excluded in quality control steps (1) and (2) described in section S2.3.2 above. Another 18 participants were excluded exclusively due to the behavioural performance criteria in step (3).

Despite the lowered relative masking threshold (from SPM default 0.8 to 0.4)[33], signal was absent in a small caudal area of the amygdala ROI for some participants, and hence this area was not included in group-level analyses. Please see Figure S1 for an illustration of the amygdala ROI. Please see Figure S2 for an illustration of an illustration of the caudal amygdala ROI where signal was absent.

#### *S3.2 Behavioural Measure Associations*

##### S3.2.1 LMDD Associations with Behavioural Measures

RTs were lower for fearful-face condition compared to neutral-face condition for control participants (mean neutral-face RT 1027 ms, mean fearful-face RT 968 ms, $t(663) = 15.635,\ P < 0.00001$). This was similar for LMDD cases (mean neutral-face RT 1029 ms, mean fearful-face RT 972 ms, $t(289) = 10.539,\ P < 0.00001$). Gender identification accuracy was slightly higher for fearful-face condition compared to neutral-face condition for control participants (mean neutral-face accuracy 97.43%, mean fearful-face accuracy 98.18%, $t(663) = 4.6673,\ P < 0.00001$), but not for participants with LMDD (mean neutral-face accuracy 97.78%, mean fearful-face accuracy 98.16%, $t(289) = 1.456,\ P = 0.1385$).

Linear regression analyses revealed that there were no significant differences between control and LMDD participants in RTs (combined conditions $P = 0.9184$, neutral-face condition $P = 0.8577$, fearful-face condition $P = 0.9840$), and in gender identification accuracies (combined conditions $P = 0.6996$, neutral-face condition $P = 0.5304$, fearful-face condition $P = 0.9857$). No significant associations of LMDD were also revealed for the difference in RT between fearful-face and neutral-face conditions ($P = 0.6967$), or for fractions of missed trials ($P = 0.2689$).

###### S3.2.2 QIDS Score Associations with Behavioural Measures

Higher severity of depression (QIDS score) was associated with higher RTs in response to both neutral and fearful face stimuli (combined conditions $\beta = 0.0664,\ P = 0.0272$, neutral-face condition $\beta = 0.0654,\ P = 0.0296$, fearful-face condition $\beta = 0.0648,\ P = 0.0323$). QIDS scores were not significantly associated with gender identification accuracy for either the entire task ($P = 0.0272$), or neutral-face and fearful-face conditions separately (respectively $P = 0.7088$ and $P = 0.2753$). QIDS scores were also not significantly associated with fractions of missed trials ($P = 0.9800$).

##### *S3.3 Brain Activation Associations*

###### S3.3.1 Group-level Brain Activations

In the entire sample ($N = 954$ participants), whole-brain analyses revealed extensive activations across the entire brain (covering frontal, occipital, temporal and parietal lobes) for both Neutral > Baseline and Fearful > Baseline contrasts with whole-brain FWE correction at $P_{FWE} < 0.05$ significance level, with strongest activations in the visual cortex. Activations were seen in bilateral occipital (MNI -14 -100 6 and 20 -96 14), fusiform (MNI -44 -56 -20 and 42 -52 -22), paracentral (MNI -6 10 52 and 6 14 52), precentral (MNI -44 0 56 and 44 2 56), prefrontal (MNI -38 20 26 and 40 42 30) and right parietal (MNI 32 -56 54) cortices (Tables S3-S4 and Figure S3). Please see Figure S3 and Tables S3-S4 for illustration and details of whole-brain activations in response to face stimuli.

Across the entire sample, analyses with SVC also revealed activations in right amygdala and in bilateral DLPFC. Mean bilateral amygdala activations, derived with the MarsBaR toolkit, were higher in the fearful-face compared to neutral-face condition, for both controls ($t(663) = 2.0051,\ P = 0.0454$) and LMDD cases ($t(289) = 2.5341,\ P = 0.0118$).

For Neutral > Fearful contrast, there were activations in bilateral occipital cortex (MNI -8 -80 44 and 20 -74 52, cluster-level $P_{FWE} < 0.001,\ k_E = 442$), right parietal (MNI 42 -34 40, cluster-level $P_{FWE} < 0.001,\ k_E = 350$), right paracentral (MNI 30 2 66, cluster-level $P_{FWE} < 0.001,\ k_E = 112$), and left DLPFC (MNI -42 38 26, cluster-level $P_{FWE} < 0.001,\ k_E = 114$). For Fearful > Neutral contrast there were activations in bilateral occipital cortex (MNI 30 -92 -2, cluster-level $P_{FWE} < 0.001,\ k_E = 475$ and MNI -34 -90 -10, cluster-level $P_{FWE} < 0.001,\ k_E = 509$) and close to right DLPFC (MNI 50 32 6, cluster-level $P_{FWE} = 0.003,\ k_E = 21$). Please see Tables S5-S6 for further details of activations related to Neutral > Fearful and Fearful > Neutral contrasts.

SVC analyses with the amygdala ROI revealed significant activations in right but not left amygdala (MNI 18 -8 -20) for Neutral > Baseline ($P_{FWE} = 0.012,\ k_E = 22$) and Fearful > Baseline ($P_{FWE} = 0.024,\ k_E = 8$) contrasts. No significant activations were revealed for Neutral > Fearful and Fearful > Neutral contrasts.

Analyses with DLPFC SVC revealed activations in bilateral DLPFC for Neutral > Baseline contrast (peaks at MNI 52 14 30, cluster-level $P_{FWE} < 0.001,\ k_E = 2232$ and MNI -52 4 42, cluster-level $P_{FWE} < 0.001,\ k_E = 1282$), for Fearful > Baseline contrast (peaks at MNI 52 14 30, cluster-level $P_{FWE} < 0.001,\ k_E = 2284$ and MNI -46 4 42, cluster-level $P_{FWE} < 0.001$, $k_E = 1271$), and for Neutral > Fearful contrast (MNI -42 38 26, $P_{FWE} < 0.001$, $k_E = 209$ and MNI 32 38 38, $P_{FWE} = 0.001$, $k_E = 67$). For Fearful > Neutral contrast there were activations in right but not left DLPFC (MNI 50 32 6, $P_{FWE} = 0.001$, $k_E = 47$).

###### S3.3.2 LMDD Associations with Brain Activations

Please see the results section of the main text for description of the differences in brain activation in LMDD cases, revealed with whole-brain and SVC analyses. Please see Tables S7-S8, Figure S4 and Tables S9-S10, Figure S5 for details and illustration of increased whole-brain activations in LMDD compared to controls, respectively in Neutral > Baseline and Fearful > Baseline contrasts, with and without correction for antidepressant medication. Please see Tables S11-S12, Figure S6 and Tables S13-S14, Figure S7 for details and illustration of increased activations in LMDD with DLPFC SVC, respectively in Neutral > Baseline and Fearful > Baseline contrasts, again with and without correction for antidepressant medication.

###### S3.3.3 QIDS Symptom Severity Associations with Brain Activations

For the entire sample ($N = 954$, both LMDD cases and controls), only a single voxel was found to be significantly negatively correlated with QIDS score for Fearful > Neutral contrast in the DLPFC SVC analysis (right hemisphere, MNI 26 40 30, $P_{FWE} = 0.038$, $k_E = 1$). No other significant associations were found.

For the LMDD sample ($N = 290$ participants, Table 1 in the main text), a cluster in the right visual cortex was found to be significantly positively correlated with QIDS for Neutral > Baseline contrast within the exploratory whole-brain analysis (MNI 16 -88 16, cluster-level $P_{FWE} = 0.047$, $k_E = 55$). No other significant associations were found.

For the control sample only ( $N = 664$ participants, Table 1 in the main text), two voxels were found to be significantly negatively correlated with QIDS score in the left amygdala for Fearful > Baseline contrast within the amygdala SVC analysis (MNI -22 -10 -22, $P_{FWE} = 0.038$, $k_E = 2$ ). Two more voxels were found to be negatively correlated with QIDS for Fearful > Baseline in the DLPFC SVC analysis (MNI 26 48 34 and -24 46 38, $P_{FWE} = 0.038$ ). In control participants there were also significant negative correlations of mean bilateral amygdala activations with QIDS scores (neutral-face condition $\beta = -0.0819$, $P = 0.0337$, fearful-face condition $\beta = -0.0898$, $P = 0.0193$ ). No other significant associations were found.

##### *S3.4 Effective Connectivity Results*

###### S3.4.1 Participant Sample in Effective Connectivity Analyses

Of $N = 954$ participants passing quality control (section S2.3.2 above), $N = 688$ were included in the DCM analyses (please see Table 2 in the main text for sample characteristics). $N = 185$ participants were excluded because they did not have sufficient activation in at least one of the three specified DCM ROIs (section S2.6.2 above). Further $N = 81$ participants had less than 10% of the variance of time-series data explained after DCM model fitting, and were excluded following recommendations from the DCM authors.[34] Mean explained variance across the $N = 688$ included participants ( $N = 474$ controls, $N = 214$ LMDD cases) was 32.07%. There are generally no strict criteria for optimal explained variance in DCM studies,[23,34] but this result is consistent with previous effective connectivity work in our lab.[33] The proportion of participants included in the DCM analysis (72.12%) was higher than in the previous work (34.74%)[33] because ROI definition was tailored for each participant based on their individual peak activation coordinates (section S2.6.2).

Participants excluded from the DCM analyses ( $N = 266$ ) were not significantly different from those included with regard to age, sex, lifetime or current MDD prevalence, antidepressant medication prevalence, QIDS scores or within-scanner task performance (Table S2). Excluded participants were more likely to be from Aberdeen rather than Dundee site, and on average had higher fractions of artifact EPI volumes detected with ArtRepair ( $t(445) = 2.32, P = 0.0206$ ). This indicates that participants were excluded from DCM analyses largely on the basis of scan quality but not clinical or demographic characteristics.

###### S3.4.2 LMDD Associations with Effective Connectivity

Please see the results section of the main text for a description of results related to changes in effective connectivity in LMDD. Tables S15-S16 provide DCM parameter estimates and estimate probabilities for full DCM model (both endogenous connections and modulatory inputs estimated), respectively with and without antidepressant medication included as a covariate. When DCM was estimated with only modulatory input parameters ('B' matrix), probability of association of LMDD with negative modulation of the Amygdala → DLPFC connection by fearful-face condition was reduced from 0.818 to 0.614. Probability of association of LMDD with reduced negative modulation of the V1 → DLPFC connection was reduced from 0.579 to zero. This was likely due to the lower flexibility of the model when the endogenous connection parameters ('A' matrix) are fixed. Tables S17-S18 provide DCM parameter estimates and estimate probabilities for the DCM model with only endogenous connections estimated ('A' matrix, 9 parameters), respectively with and without antidepressant medication covariate. Tables S19-S20 provide DCM parameter estimates and estimate probabilities for the DCM model with only modulatory inputs estimated ('B' matrix, 18 parameters), again with and without antidepressant medication covariate.

###### S3.4.3 QIDS Symptom Severity Associations with Effective Connectivity

No significant associations of QIDS scores were found in the full DCM model (when LMDD status was replaced with QIDS). Analysis with separate estimation of only endogenous connectivity parameters (‘A’ matrix) also did not reveal any associations of QIDS. Analysis with separate estimation of only modulatory input parameters parameters (‘B’ matrix) revealed that higher QIDS scores were related to: 1) Small increase in modulation of inhibitory self-connection of the DLPFC by the neutral-face condition (estimate 0.02, probability 0.61), and 2) Small increase in modulation of V1 → Amygdala connection by the fearful-face condition (estimate 0.012, probability 0.64). These results indicate that higher QIDS scores are related to increased self-inhibition of the DLPFC in neutral-face condition, and increased connectivity from V1 to amygdala in fearful-face condition. Tables S21-S23 provide DCM parameter estimates and estimate probabilities respectively for the full DCM model, DCM model with only endogenous connections (‘A’ matrix), and DCM model with only modulatory inputs (‘B’ matrix), with QIDS scores as the main predictor of interest.

**Table S1 | Summary of studies investigating changes in brain activation in response to fearful face stimuli in depression (1 / 10)**

| Authors | Year | Journal | Diagnoses | Medications | Sample size | Paradigm | Analysis | Results | Result regions | Result conditions |
|---|---|---|---|---|---|---|---|---|---|---|
| *Wackerhagen et al.*[10] | 2020 | Psychological Medicine | Current MDD<br>MDD relatives | Medicated | Controls – 106<br>Depressed – 48<br>Relatives – 49 | Emotional face matching | 1) Whole-brain<br>2) Amygdala ROI gPPI connectivity from bilateral amygdala to rest of the brain | Negative correlation of HAM-D with activations in MDD | - Left superior frontal gyrus | Combined fearful and anger faces vs. shapes |
| | | | | | | | | Decreased connectivity in MDD compared to controls and relatives | - Amygdala and right medial prefrontal cortex, posterior cingulate cortex, parietal cortex | Combined conditions |
| | | | | | | | | | - Amygdala and right middle frontal gyrus | Combined fearful and anger faces vs. shapes |
| | | | | | | | | Decreased connectivity in MDD and relatives compared to controls | - Amygdala and superior parietal cortex / right superior frontal gyrus | Combined conditions |
| | | | | | | | | | - Amygdala and right fusiform gyrus | Combined fearful and anger faces vs. shapes |
| *Alders et al.*[35] | 2019 | Journal of Affective Disorders | Current MDD | Medication-free | Controls – 30<br>Depressed – 48 | Face emotion recognition with emotion word distractions | Whole-brain | Decreased activations in MDD compared to controls in all conditions | - Right inferior temporal gyrus<br>- Lateral occipital cortex<br>- Occipital fusiform gyrus | - All trials<br>- Congruent trials<br>- Incongruent trials<br>- Inc. vs. con. contrast |

**Table S1 | Summary of studies investigating changes in brain activation in response to fearful face stimuli in depression (continued 2 / 10)**

| Authors | Year | Journal | Diagnoses | Medications | Sample size | Paradigm | Analysis | Results | Result regions | Result conditions |
|---|---|---|---|---|---|---|---|---|---|---|
| *Korgaonkar et al.*[1] | 2019 | Biological Psychiatry: CNNI | Remitted MDD<br>Remitted bipolar | Medicated | Controls – 25<br>Remitted depressed – 25<br>Remitted bipolar – 31 | Emotional face viewing with subliminal and supraliminal stimuli | 1) Whole-brain<br>2) Amygdala ROI<br>3) gPPI connectivity from left amygdala to sgACC, pgACC, hippocampus, insula | Increased activation in remitted MDD compared to controls | - Left amygdala | Subliminal combined fear / anger / disgust faces |
| | | | | | | | | Decreased activation in remitted bipolar compared to remitted MDD | | - Supraliminal disgust faces<br>- Supraliminal fear faces<br>- Subliminal neutral faces<br>- Subliminal sad faces<br>- Subliminal happy faces<br>- Subliminal threat faces |
| | | | | | | | | Increased connectivity in remitted MDD compared to controls | - Left amygdala and hippocampus | Supraliminal sad faces |
| | | | | | | | | | - Left amygdala and medial OFC | Supraliminal happy faces |
| | | | | | | | | Decreased connectivity in remitted bipolar compared to remitted MDD | - Left amygdala with hippocampus | - Supraliminal sad faces<br>- Supraliminal neutral faces<br>- Subliminal neutral faces |
| | | | | | | | | | - Left amygdala with right amygdala | - Supraliminal neutral faces<br>- Subliminal sad faces |
| | | | | | | | | | - Left amygdala with medial OFC / left putamen / left caudate | - Supraliminal happy faces<br>- Subliminal happy faces |
| | | | | | | | | | - Left amygdala with insula<br>- Left amygdala with hippocampus | - Subliminal anger faces<br>- Subliminal disgust faces |
| | | | | | | | | Increased connectivity in remitted bipolar compared to remitted MDD | - Left amygdala with insula | - Supraliminal sad faces<br>- Subliminal sad faces |
| *Li et al.*[36] | 2018 | BMC Psychiatry | Current MDD | Not medicated | Controls – 32<br>Depressed – 36 | Gender recognition of emotional faces | 1) Whole-brain<br>2) Spatial ICA | No group differences | - | - |

**Table S1 | Summary of studies investigating changes in brain activation in response to fearful face stimuli in depression (continued 3 / 10)**

| Authors | Year | Journal | Diagnoses | Medications | Sample size | Paradigm | Analysis | Results | Result regions | Result conditions |
|---|---|---|---|---|---|---|---|---|---|---|
| *Stange et al.*[37] | 2018 | Journal of Affective Disorders | Remitted MDD | Medication-free | Controls – 33<br>Remitted – 43 | Face emotion recognition | 1) Cognitive control network<br>2) Salience and emotion network | Increased activation in remitted MDD compared to controls | Cognitive control network | - Fearful faces<br>- Sad faces |
| | | | | | | | | Increased activation extent in remitted MDD compared to controls | Salience and emotion network | Sad faces |
| *Bürger et al.*[8] | 2017 | Neuropsycho-pharmacology | Current MDD<br>Bipolar | Medicated | Controls – 36<br>Depressed – 36<br>Bipolar – 36 | Emotional faces matching | 1) Whole-brain<br>2) Amygdala ROI<br>3) ACC ROI | Decreased activations in MDD compared to controls or bipolar | - Anterior cingulate cortex<br>- Middle cingulate cortex<br>- Superior frontal gyrus | Fearful faces |
| | | | | | | | | | - Basal ganglia - Insula<br>- Amygdala - Hippocampus<br>- Cingulate cortex | Happy faces |
| | | | | | | | | Positive correlation of BDI with activations | - Anterior cingulate cortex | Fearful faces |
| | | | | | | | | Negative correlation of HAM-D with activations | | - Fearful faces<br>- Happy faces |
| | | | | | | | | Negative correlation of HAM-A with activations | | Angry faces |

**Table S1 | Summary of studies investigating changes in brain activation in response to fearful face stimuli in depression (continued 4 / 10)**

| Authors | Year | Journal | Diagnoses | Medications | Sample size | Paradigm | Analysis | Results | Result regions | Result conditions |
|---|---|---|---|---|---|---|---|---|---|---|
| *Luo et al.*[13] | 2017 | Human Brain Mapping | None | Not medicated | Controls – 92 | Face emotion recognition<br>Emotional faces matching | 1) Whole-brain<br>2) gPPI connectivity from left globus pallidus / left caudate to rest of the brain | Positive correlation of BDI with activations | - Left caudate<br>- Right globus pallidus<br>- Left inferior parietal cortex<br>- ACC / PCC<br>- Inferior frontal gyrus<br>- Left medial PFC<br>- Right hippocampus<br>- Fusiform gyrus<br>- Right precentral cortex<br>- Left postcentral cortex<br>- Right precuneus<br>- Middle temporal cortex<br>- Left lingual gyrus<br>- Right superior frontal cortex<br>- Left middle frontal gyrus<br>- Left SMA<br>- Cerebellum | Fearful faces recognition |
| | | | | | | | | Negative correlation of BDI with connectivity | - Left putamen to right superior temporal gyrus / right amygdala | |
| *MacNamara et al.*[38] | 2017 | Depression and Anxiety | Current MDD<br>Current SAD<br>Current anxiety | Medication-free | Controls – 57<br>Depressed – 43<br>Affective – 79<br>Anxiety – 20 | Face emotional expression matching | Whole-brain | Increased activation in combined patients compared to controls | - Occipital / lingual gyrus | - Anger faces<br>- Fearful faces |
| | | | | | | | Whole-brain correlation | Positive correlation of HAM-A with activations | - Insula<br>- ACC / MCC<br>- Right DLPFC | Anger faces |
| | | | | | | | | Negative correlation of HAM-D with activations | - Right DLPFC | |
| *Powers et al.*[12] | 2017 | Social Cognitive and Affective Neuroscience | None | Medication-free | Controls – 92 | Emotional faces viewing | DMPFC ROI<br>VMPFC ROI<br>Middle temporal ROI<br>Inferior frontal ROI | Positive correlation of BDI with activation | - Dorsomedial PFC | Fearful faces vs. neutral faces contrast |
| *Ferri et al.*[39] | 2017 | Cognitive, Affective, & Behavioral Neuroscience | Current MDD (treatment resistant) | Medicated | Controls – 37<br>Depressed – 80 | 1) Face emotion recognition<br>2) Emotional faces viewing<br>3) Gender recognition of emotional faces | Amygdala ROI | Decreased activation in TRD compared to controls | - Bilateral amygdala | Face emotion recognition (non-specific emotions) |
| | | | | | | | | *Negative correlation of HAM-D with activation in TRD* | | |

**Table S1 | Summary of studies investigating changes in brain activation in response to fearful face stimuli in depression (continued 5 / 10)**

| Authors | Year | Journal | Diagnoses | Medications | Sample size | Paradigm | Analysis | Results | Result regions | Result conditions |
|---|---|---|---|---|---|---|---|---|---|---|
| *Müller et al.*[40] | 2014 | Social Cognitive and Affective Neuroscience | Current MDD | Medicated | Controls – 21<br>Depressed – 21 | Emotional face rating with emotional sound distractions | Whole-brain | Increased activation in MDD compared to controls | - Left fusiform gyrus | All trials |
| | | | | | | | | | - Left inferior parietal cortex<br>- Left inferior frontal gyrus | Happy faces with congruent distractions |
| | | | | | | | | | - Right superior temporal cortex<br>- Posterior superior temporal<br>- Middle cingulate cortex | Neutral faces with emotional distractions |
| | | | | | | | | Negative correlation of BDI with activation in MDD | - Right superior temporal cortex | Emotional target and distraction vs. emotional target or distraction contrast |
| *Fournier et al.*[41] | 2013 | Bipolar Disorders | Current MDD<br>Bipolar | Medicated | Controls – 29<br>Depressed – 30<br>Bipolar – 22 | Colour recognition with emotional face distractions | 1) Whole-brain<br>2) Amygdala ROI | Increased activation in current MDD compared to controls or bipolar | - Left amygdala | Anger faces |
| | | | | | | | | Increased activation in current MDD compared to controls | - Right ACC<br>- Temporal cortex<br>- Parietal cortex | Anger faces |
| | | | | | | | | | - Left temporal cortex<br>- Right parietal cortex | Happy faces |
| | | | | | | | | Increased activation in current MDD compared to bipolar | - Right ACC<br>- Right parietal cortex<br>- Temporal cortex<br>- Parietal cortex<br>- Occipital cortex<br>- Insula | - Anger faces<br>- Fearful faces<br>- Happy faces |
| *Fournier et al.*[42] | 2013 | Psychological Medicine | Current MDD | Medicated | Controls – 28<br>Depressed – 26 | Colour recognition with emotional face distractions | 1) Whole-brain<br>2) Amygdala ROI | Increased activation in current MDD compared to controls | - Right amygdala<br>- Occipitotemporal cortex<br>- Frontal cortex<br>- Parietal cortex | - Anger faces<br>- Happy faces |

**Table S1 | Summary of studies investigating changes in brain activation in response to fearful face stimuli in depression (continued 6 / 10)**

| Authors | Year | Journal | Diagnoses | Medications | Sample size | Paradigm | Analysis | Results | Result regions | Result conditions |
|---|---|---|---|---|---|---|---|---|---|---|
| *Greening et al.*[2] | 2013 | Journal of Affective Disorders | Current MDD | Medication-free | Controls – 18<br>Depressed – 18 | Face emotion recognition with emotion face distractions | 1) Whole-brain<br>2) Amygdala ROI | Increased activation in current MDD compared to controls | - Right amygdala | Fearful face target trials |
| | | | | | | | | Decreased activation in current MDD compared to controls | - Right amygdala | Happy face target trials |
| | | | | | | | | | - Left amygdala<br>- Left parahippocampal gyrus | Happy face distractions |
| | | | | | | | | | - Dorsomedial PFC<br>- Ventrolateral PFC | Negative distractions |
| *Kong et al.*[43] | 2013 | Journal of Psychiatry and Neuroscience | Current MDD | Not medicated | Controls – 30<br>Depressed – 28 | Gender recognition of emotional faces | 1) Whole-brain to amygdala connectivity<br>2) Amygdala to PFC ROI connectivity | Decreased functional connectivity in MDD compared to controls | - Left rostral PFC and amygdala | Fearful faces |
| *Arnone et al.*[44] | 2012 | American Journal of Psychiatry | Current MDD<br>Remitted MDD | Medication-free | Controls – 54<br>Depressed – 38<br>Remitted – 24 | Gender recognition of emotional faces | Whole-brain<br>Amygdala ROI | Increased activation in current MDD compared to controls and remitted MDD | - Left / right amygdala | 1) Sad faces<br>2) Sad faces vs. neutral faces contrast |
| | | | | | | | | Negative correlation of change in MARDS with activation in MDD | - Parahippocampal gyrus | Sad faces |
| *Kerestes et al.*[9] | 2012 | Psychiatry Research Neuromaging | Remitted MDD | Medication-free | Controls – 20<br>Remitted – 19 | Gender recognition of emotional faces | Whole-brain<br>DLPFC ROI<br>VLPFC ROI<br>OFC ROI<br>Subgenual ACC ROI<br>Amygdala ROI<br>Ventral striatum ROI | Decreased activation in remitted MDD compared to controls | - Left DLPFC | 1) Fearful faces<br>2) Fearful faces vs. neutral faces contrast |
| | | | | | | | | | - Left OFC | Fearful faces vs. neutral faces contrast |
| | | | | | | | | *Positive correlation of euthymia duration with activation in remitted MDD* | - Right DLPFC | Fearful faces vs. neutral faces contrast |

**Table S1 | Summary of studies investigating changes in brain activation in response to fearful face stimuli in depression (continued 7 / 10)**

| Authors | Year | Journal | Diagnoses | Medications | Sample size | Paradigm | Analysis | Results | Result regions | Result conditions |
|---|---|---|---|---|---|---|---|---|---|---|
| *Ruhe et al.*[3] | 2012 | Journal of Clinical Psychiatry | Current MDD | Medicated | Controls – 21<br>Depressed – 20 | Gender recognition of emotional faces | 1) Whole brain<br>2) Amygdala ROI | *Increased activation in MDD compared to controls* | - Right amygdala<br>- Left insula | Combined fearful and anger faces |
| | | | | | | | | | - Left subthalamic nucleus | Happy faces |
| | | | | | | | | *Decreased activation in MDD compared to controls* | - VLPFC<br>- Left DMPFC<br>- Right DLPFC<br>- Left cerebellum<br>- Left fusiform gyrus<br>- Left posterior cingulate cortex<br>- Right superior temporal cortex | Combined fearful and anger faces |
| | | | | | | | | | - Right VLPFC<br>- Right precentral cortex<br>- Right superior temporal cortex<br>- Left fusiform gyrus | Happy faces |
| *Almeida et al.*[45] | 2011 | Frontiers in Psychiatry | Current MDD | Medicated | Controls – 19<br>Depressed – 19 | Color recognition with emotional face distractions | DCM with ventromedial PFC / subgenual ACC / amygdala | Increased excitatory connection in current MDD compared to controls | - Left sgACC to left amygdala | Fearful faces |
| | | | | | | | | Increased inhibitory connections in female MDD compared to female controls | - Left vmPFC to left amygdala<br>- Left sgACC to left amygdala | Happy faces |
| | | | | | | | | Decreased excitatory connection in female MDD compared to female controls | - Left vmPFC to left sgACC | |
| | | | | | | | | Negative correlation of HAM-D with connectivity in controls | - Left sgACC to left amygdala | |
| *Demenescu et al.*[46] | 2011 | Psychological Medicine | Current MDD<br>Remitted MDD<br>Current anxiety<br>Co-morbidity (anx. and MDD) | Medicated | Controls – 56<br>Depressed or Remitted – 59<br>Anxiety – 57<br>Co-morbid – 66 | Gender recognition of emotional faces | 1) Whole-brain<br>2) Amygdala ROI | Increased activation in current / remitted MDD compared to controls | - Superior frontal gyrus<br>- Middle frontal gyrus | Happy faces |
| | | | | | | | | Decreased activation in anxiety compared to controls | - Right lentiform nucleus | |
| | | | | | | | | Positive correlation of MADRS with activation in current MDD | - Left fusiform gyrus | - Angry faces<br>- Fearful faces |

**Table S1 | Summary of studies investigating changes in brain activation in response to fearful face stimuli in depression (continued 8 / 10)**

| Authors | Year | Journal | Diagnoses | Medications | Sample size | Paradigm | Analysis | Results | Result regions | Result conditions |
|---|---|---|---|---|---|---|---|---|---|---|
| *Matthews et al.*[4] | 2011 | NeuroImage | Current MDD | Partly medicated | Controls – 11<br>Depressed – 11 | Emotional faces matching | 1) Whole-brain<br>2) Amygdala ROI | Increased activation in MDD compared to controls | - Amygdala<br>- Right thalamus<br>- Right cerebellum<br>- Right middle temporal cortex | Fearful faces |
| | | | | | | | | Decreased activation in MDD compared to controls | - Right inferior parietal cortex<br>- Left postcentral cortex<br>- Right paracentral cortex<br>- Left middle frontal cortex<br>- Right cuneus<br>- Left DLPFC | |
| *Thomas et al.*[47] | 2011 | Psychological Medicine | Remitted MDD | Medication-free | Controls – 35<br>Remitted – 28 | Gender recognition of emotional faces | - Amygdala ROI<br>- Hippocampus ROI<br>- Parahipp. ROI<br>- Globus pallidus ROI<br>- Thalamus ROI<br>- Caudate ROI<br>- Putamen ROI<br>- Insula ROI<br>- Fusiform ROI<br>- PFC ROI<br>- OFC ROI<br>- Cingulate ROI | Decreased activation in remitted MDD compared to controls | - Right insula<br>- Right fusiform gyrus<br>- Rright putamen<br>- Hippocampus | Sad faces vs. neutral faces contrast |
| | | | | | | | | | - Left hippocampus<br>- Left pars opercularis | Fearful faces vs. neutral faces contrast |
| | | | | | | | | Positive correlation of RRS with activation in remitted MDD | - Right insula<br>- MCC | Sad faces |
| | | | | | | | | | - Left precentral gyrus<br>- Right parahippocampal gyrus<br>- Left insula<br>- Right caudate | Fearful faces |
| | | | | | | | | Negative correlation of RRS with activation in remitted MDD | - Medial frontal gyrus<br>- Right inferior frontal gyrus<br>- Hippocampus<br>- Parahippocampal gyrus<br>- Left thalamus<br>- Left globus pallidus | Happy faces |

**Table S1 | Summary of studies investigating changes in brain activation in response to fearful face stimuli in depression (continued 9 / 10)**

| Authors | Year | Journal | Diagnoses | Medications | Sample size | Paradigm | Analysis | Results | Result regions | Result conditions |
|---|---|---|---|---|---|---|---|---|---|---|
| *Van Wingen et al.*[48] | 2011 | Psychological Medicine | Current MDD<br>Remitted MDD | Not medicated or medication-free | Controls – 30<br>Depressed – 18<br>Remitted – 18 | 1) Emotional face expressions matching<br>2) Face emotion recognition | 1) Whole-brain<br>2) Amygdala ROI | Increased activation in current MDD compared to controls / remitted MDD | - Right intraparietal cortex<br>- Left inferior frontal cortex<br>- Right inferior occipital cortex<br>- Left middle occipital cortex<br>- Right putamen<br>- Right insula<br>- Right amygdala | Emotion recognition |
| | | | | | | | | Decreased activation in current MDD compared to controls / remitted MDD | - Left precuneus<br>- ACC | Emotional expression matching |
| *Norbury et al.*[11] | 2010 | Psychological Medicine | Remitted MDD | Medication-free | Controls – 21<br>Remitted – 16 | Emotional face expressions matching | 1) Whole-brain<br>2) Amygdala ROI | Increased activation in remitted MDD compared to controls | - Left DLPFC<br>- Right DLPFC<br>- Right caudate | Fearful faces |
| | | | | | | | | Decreased activation in remitted MDD compared to controls | - Left DLPFC | Happy faces |
| *Surguladze et al.*[49] | 2010 | Journal of Psychiatric Research | Current MDD | Medicated | Controls – 9<br>Depressed – 9 | Gender recognition of emotional faces | Whole-brain | Increased activation in MDD compared to controls | - Left insula<br>- Left OFC<br>- Middle temporal cortex | Disgust faces |
| | | | | | | | | Decreased activation in MDD compared to controls | - Left inferior parietal cortex | Fearful faces |
| *Moses-Kolko et al.*[7(p)] | 2010 | American Journal of Psychiatry | Current MDD | Medication-free | Controls – 16<br>Depressed – 14 | Emotional faces matching | 1) Amygdala ROI<br>2) DMPFC ROI<br>3) Granger causality between DMPFC and amygdala activation | Decreased activation in MDD compared to controls | - Left DMPFC | Combined fear and anger faces |
| | | | | | | | | Increased activation in MDD compared to controls | - Left DMPFC | Shape trials |
| | | | | | | | | Negative correlation of with HAM-D scores in MDD | - Left amygdala | Combined fear and anger faces |
| | | | | | | | | Absent effective connectivity in MDD but not controls | - Left DMPFC to left amygdala | |

**Table S1 | Summary of studies investigating changes in brain activation in response to fearful face stimuli in depression (continued 10 / 10)**

| Authors | Year | Journal | Diagnoses | Medications | Sample size | Paradigm | Analysis | Results | Result regions | Result conditions |
|---|---|---|---|---|---|---|---|---|---|---|
| *Townsend et al.*[50] | 2010 | Psychiatry Research Neuromaging | Current MDD | Medication-free | Controls – 15<br>Depressed – 15 | Emotional face expressions matching | 1) Whole-brain<br>2) Amygdala ROI<br>3) OFC ROI | Decreased activation in MDD compared to controls | - Insula<br>- Inferior temporal gyrus<br>- Middle temporal gyrus<br>- Hippocampus<br>- Putamen<br>- Occipital gyrus<br>- Fusiform gyrus<br>- Cerebellum | All trials |
| *Peluso et al.*[5] | 2009 | Psychiatry Research Neuromaging | Current MDD | Medication-free | Controls – 15<br>Depressed – 14 | 1) Face emotion recognition<br>2) Emotional faces matching | 1) Whole-brain<br>2) Amygdala ROI | Increased activation in MDD compared to controls | - Amygdala | Combined fear and anger faces |
| | | | | | | | | Positive correlation of BDI with activation in MDD | | |
| *Matthews et al.*[51] | 2008 | Journal of Affective Disorders | Current MDD | Medication-free | Controls – 16<br>Depressed – 15 | Emotional face matching | 1) Extended amygdala ROI<br>2) Functional connectivity between extended amygdala and ACC | Increased activation in MDD compared to controls | - Extended amygdala | Face matching vs. shape matching con. |
| | | | | | | | | Increased functional connectivity in MDD compared to controls | - Bilateral extended amygdala and sgACC | Combined face matching and shape matching task conditions |
| | | | | | | | | Decreased functional connectivity in MDD compared to controls | - Bilateral extended amygdala and dorsal ACC | |
| | | | | | | | | Negative correlation of BDI with connectivity | | |
| *Fales et al.*[6] | 2008 | Biological Psychiatry | Current MDD | Medication-free | Controls – 24<br>Depressed – 27 | Face matching and neutral image matching with emotional faces as either targets or distractions | 1) Whole-brain<br>2) Amygdala ROI<br>3) Subgenual ACC ROI<br>4) Pregenual ACC ROI<br>5) Rostral ACC ROI<br>6) Dorsal ACC ROI<br>7) DLPFC ROIs | Increased activation in MDD compared to controls | - Left amygdala | Ignore fear faces vs. ignore neutral faces contrast |
| | | | | | | | | | - Pregenual ACC | Ignore faces trials |
| | | | | | | | | | - Subgenual ACC | All trials |
| | | | | | | | | Decreased activation in MDD compared to controls | - Left amygdala | Attend fear faces vs. attend neutral faces contrast |
| | | | | | | | | | - Right DLPFC | Ignore fear faces vs. ignore neutral faces contrast |
| | | | | | | | | | - Pregenual ACC | Attend faces trials |
| | | | | | | | | | - Rostral ACC<br>- Dorsal ACC | All trials |

**Table S2**

**Summary demographic characteristics of the participants included and excluded from the effective connectivity (DCM) analyses**

| | **Included in DCM** | **Excluded from DCM** | ***P* value** |
|---|---|---|---|
| Size (*N*) | 688 | 266 | - |
| Sex (male / female) | 283 / 405 | 92 / 174 | n.s. |
| Age (years) | 59.07 (10.08) | 58.22 (10.50) | n.s. |
| Site (Aberdeen / Dundee) | 320 / 368 | 170 / 96 | < 0.001 |
| Lifetime MDD (*N*) | 214 (31.1%) | 76 (28.6%) | n.s. |
| Current MDD (*N*) | 32 (4.7%) | 9 (3.4%) | n.s. |
| Medicated (*N*) | 100 (14.5%) | 32 (12.0%) | n.s. |
| QIDS (score) | 4.52 (3.59) | 4.67 (3.57) | n.s. |
| Artifact volume fraction | 1.08% (1.88%) | 1.42% (2.06%) | 0.0206 |
| Normalisation issues (*N*) | 59 (8.6%) | 17 (6.4%) | n.s. |
| Task accuracy | 96.52% (12.23%) | 95.85% (13.49%) | n.s. |
| Missed trials fraction | 0.75% (2.26%) | 0.92% (2.41%) | n.s. |

***Note***: Participants were considered medicated if they had an antidepressant prescription at the time of the scan. Standard deviations or percentage fractions from overall sample are in parentheses where relevant. *P*-values defined according to two-sample t-tests (continuous measures) or chi-squared tests (proportions). Significant differences between DCM included and excluded participants highlighted in light blue.

**Table S3**

**Details of significant brain activations in response to *neutral* faces (Neutral > Baseline contrast) in complete sample (whole-brain FWE with significance level $P_{FWE} < 0.05$)**

| set-level | | cluster-level | | | | peak-level | | | | | mm mm mm | | |
|---|---|---|---|---|---|---|---|---|---|---|---|---|---|
| $p$ | $c$ | $p_{FWE-corr}$ | $q_{FDR-corr}$ | $k_E$ | $p_{uncorr}$ | $p_{FWE-corr}$ | $q_{FDR-corr}$ | $T$ | $(Z_E)$ | $p_{uncorr}$ | | | |
| 0.000 | 15 | 0.000 | 0.000 | 9641 | 0.000 | 0.000 | 0.000 | 31.74 | Inf | 0.000 | -12 | -100 | 6 |
| | | | | | | 0.000 | 0.000 | 26.90 | Inf | 0.000 | 20 | -96 | 14 |
| | | | | | | 0.000 | 0.000 | 22.62 | Inf | 0.000 | 12 | -92 | -6 |
| | | 0.000 | 0.000 | 13814 | 0.000 | 0.000 | 0.000 | 15.80 | Inf | 0.000 | 52 | 14 | 30 |
| | | | | | | 0.000 | 0.000 | 15.35 | Inf | 0.000 | -52 | -24 | 50 |
| | | | | | | 0.000 | 0.000 | 15.35 | Inf | 0.000 | 6 | 14 | 52 |
| | | 0.000 | 0.000 | 463 | 0.000 | 0.000 | 0.000 | 8.26 | Inf | 0.000 | -34 | 24 | -2 |
| | | | | | | 0.000 | 0.000 | 6.90 | 6.82 | 0.000 | -56 | 18 | 0 |
| | | 0.000 | 0.000 | 236 | 0.000 | 0.000 | 0.000 | 7.86 | 7.73 | 0.000 | 8 | -30 | -2 |
| | | | | | | 0.000 | 0.000 | 7.69 | 7.57 | 0.000 | 22 | -24 | -4 |
| | | 0.000 | 0.000 | 334 | 0.000 | 0.000 | 0.000 | 7.21 | 7.11 | 0.000 | 26 | -2 | 4 |
| | | | | | | 0.002 | 0.046 | 5.38 | 5.33 | 0.000 | -2 | -10 | -12 |
| | | | | | | 0.004 | 0.086 | 5.24 | 5.21 | 0.000 | 12 | -10 | -8 |
| | | 0.000 | 0.000 | 121 | 0.000 | 0.000 | 0.000 | 6.64 | 6.56 | 0.000 | -24 | -2 | 6 |
| | | 0.000 | 0.001 | 102 | 0.000 | 0.000 | 0.000 | 6.41 | 6.34 | 0.000 | -22 | -50 | 0 |
| | | 0.000 | 0.000 | 170 | 0.000 | 0.000 | 0.002 | 6.02 | 5.96 | 0.000 | 0 | -54 | -24 |
| | | | | | | 0.001 | 0.016 | 5.59 | 5.54 | 0.000 | 10 | -50 | -20 |
| | | 0.002 | 0.067 | 26 | 0.040 | 0.000 | 0.002 | 5.98 | 5.93 | 0.000 | -22 | -24 | -6 |
| | | 0.015 | 0.408 | 6 | 0.299 | 0.008 | 0.162 | 5.11 | 5.07 | 0.000 | 28 | -68 | 28 |
| | | 0.020 | 0.497 | 4 | 0.398 | 0.010 | 0.202 | 5.05 | 5.02 | 0.000 | 36 | 56 | 12 |
| | | 0.012 | 0.347 | 8 | 0.232 | 0.018 | 0.363 | 4.91 | 4.88 | 0.000 | 20 | -54 | -2 |
| | | 0.028 | 0.600 | 2 | 0.560 | 0.026 | 0.520 | 4.83 | 4.80 | 0.000 | -10 | -58 | -4 |
| | | 0.028 | 0.600 | 2 | 0.560 | 0.037 | 0.739 | 4.74 | 4.72 | 0.000 | -8 | -86 | 36 |
| | | 0.035 | 0.694 | 1 | 0.694 | 0.044 | 0.868 | 4.70 | 4.68 | 0.000 | -38 | 6 | -28 |

Height threshold: T = 4.67, p = 0.000 (0.050)
Extent threshold: k = 0 voxels
Expected voxels per cluster, <k> = 6.010
Expected number of clusters, <c> = 0.05
FWEp: 4.670, FDRp: 5.376, FWEc: 1, FDRc: 102

Degrees of freedom = [1.0, 950.0]
FWHM = 11.1 11.2 11.5 mm mm mm; 5.6 5.6 5.7 {voxels}
Volume: 1326784 = 165848 voxels = 853.2 resels
Voxel size: 2.0 2.0 2.0 mm mm mm; (resel = 178.22 voxels)

**Table S4**

**Details of significant brain activations in response to *fearful* faces (Fearful > Baseline contrast) in complete sample (whole-brain FWE with significance level $P_{FWE}$ < 0.05)**

| set-level | | cluster-level | | | | peak-level | | | | | mm mm mm | | |
|---|---|---|---|---|---|---|---|---|---|---|---|---|---|
| $p$ | $c$ | $p_{FWE-corr}$ | $q_{FDR-corr}$ | $k_E$ | $p_{uncorr}$ | $p_{FWE-corr}$ | $q_{FDR-corr}$ | $T$ | $(Z_E)$ | $p_{uncorr}$ | | | |
| 0.000 | 16 | 0.000 | 0.000 | 8882 | 0.000 | 0.000 | 0.000 | 31.47 | Inf | 0.000 | -14 | -100 | 6 |
| | | | | | | 0.000 | 0.000 | 27.31 | Inf | 0.000 | 14 | -98 | 8 |
| | | | | | | 0.000 | 0.000 | 27.02 | Inf | 0.000 | 20 | -96 | 14 |
| | | 0.000 | 0.000 | 7753 | 0.000 | 0.000 | 0.000 | 16.27 | Inf | 0.000 | 50 | 14 | 30 |
| | | | | | | 0.000 | 0.000 | 15.31 | Inf | 0.000 | 44 | 8 | 30 |
| | | | | | | 0.000 | 0.000 | 14.77 | Inf | 0.000 | 6 | 14 | 52 |
| | | 0.000 | 0.000 | 3468 | 0.000 | 0.000 | 0.000 | 14.79 | Inf | 0.000 | -52 | -24 | 52 |
| | | | | | | 0.000 | 0.000 | 14.41 | Inf | 0.000 | -44 | 0 | 56 |
| | | | | | | 0.000 | 0.000 | 13.35 | Inf | 0.000 | -32 | -2 | 66 |
| | | 0.000 | 0.000 | 406 | 0.000 | 0.000 | 0.000 | 9.82 | Inf | 0.000 | 32 | -56 | 54 |
| | | 0.000 | 0.000 | 411 | 0.000 | 0.000 | 0.000 | 8.87 | Inf | 0.000 | -34 | 24 | 0 |
| | | | | | | 0.000 | 0.004 | 5.89 | 5.83 | 0.000 | -56 | 18 | 0 |
| | | 0.000 | 0.001 | 107 | 0.000 | 0.000 | 0.000 | 6.53 | 6.46 | 0.000 | 26 | 2 | 2 |
| | | 0.000 | 0.003 | 78 | 0.001 | 0.000 | 0.000 | 6.43 | 6.36 | 0.000 | 22 | -26 | -2 |
| | | | | | | 0.000 | 0.000 | 6.31 | 6.24 | 0.000 | 8 | -30 | -2 |
| | | 0.006 | 0.158 | 15 | 0.109 | 0.001 | 0.012 | 5.66 | 5.62 | 0.000 | 60 | -16 | 20 |
| | | 0.000 | 0.010 | 56 | 0.005 | 0.002 | 0.040 | 5.41 | 5.37 | 0.000 | 0 | -54 | -24 |
| | | 0.013 | 0.351 | 7 | 0.263 | 0.003 | 0.059 | 5.33 | 5.29 | 0.000 | -24 | -24 | -6 |
| | | 0.001 | 0.039 | 34 | 0.022 | 0.003 | 0.060 | 5.32 | 5.28 | 0.000 | -26 | 2 | 0 |
| | | 0.024 | 0.576 | 3 | 0.468 | 0.004 | 0.085 | 5.24 | 5.20 | 0.000 | -8 | -90 | 30 |
| | | 0.004 | 0.119 | 19 | 0.074 | 0.005 | 0.104 | 5.19 | 5.16 | 0.000 | -22 | -52 | 0 |
| | | 0.028 | 0.598 | 2 | 0.560 | 0.007 | 0.141 | 5.13 | 5.09 | 0.000 | 10 | 30 | 64 |
| | | 0.028 | 0.598 | 2 | 0.560 | 0.010 | 0.192 | 5.06 | 5.02 | 0.000 | -58 | 16 | 4 |
| | | 0.035 | 0.694 | 1 | 0.694 | 0.027 | 0.544 | 4.81 | 4.78 | 0.000 | 14 | 28 | 64 |

Height threshold: T = 4.67, p = 0.000 (0.050)
Extent threshold: k = 0 voxels
Expected voxels per cluster, <k> = 6.030
Expected number of clusters, <c> = 0.05
FWEp: 4.669, FDRp: 5.409, FWEc: 1, FDRc: 34

Degrees of freedom = [1.0, 950.0]
FWHM = 11.2 11.2 11.4 mm mm mm; 5.6 5.6 5.7 {voxels}
Volume: 1326784 = 165848 voxels = 850.6 resels
Voxel size: 2.0 2.0 2.0 mm mm mm; (resel = 178.75 voxels)

**Table S5**

**Details of significant brain activations for Neutral > Fearful contrast in complete sample (whole-brain FWE with significance level $P_{FWE}$ < 0.05)**

| set-level | | cluster-level | | | | peak-level | | | | | mm mm mm | | |
|---|---|---|---|---|---|---|---|---|---|---|---|---|---|
| $p$ | $c$ | $p_{FWE-corr}$ | $q_{FDR-corr}$ | $k_E$ | $p_{uncorr}$ | $p_{FWE-corr}$ | $q_{FDR-corr}$ | $T$ | $(Z_E)$ | $p_{uncorr}$ | | | |
| **0.000** | **8** | **0.000** | **0.000** | **350** | **0.000** | **0.000** | **0.000** | **7.31** | **7.21** | **0.000** | **42** | **-34** | **40** |
| | | | | | | 0.032 | 0.644 | 4.77 | 4.74 | 0.000 | 46 | -22 | 32 |
| | | **0.000** | **0.000** | **442** | **0.000** | **0.000** | **0.001** | **6.50** | **6.42** | **0.000** | **-8** | **-80** | **44** |
| | | | | | | 0.000 | 0.001 | 6.40 | 6.33 | 0.000 | 20 | -74 | 52 |
| | | | | | | 0.000 | 0.006 | 6.09 | 6.03 | 0.000 | 20 | -62 | 64 |
| | | **0.000** | **0.001** | **114** | **0.000** | **0.000** | **0.006** | **6.09** | **6.03** | **0.000** | **-42** | **38** | **26** |
| | | | | | | 0.006 | 0.238 | 5.17 | 5.13 | 0.000 | -34 | 34 | 22 |
| | | **0.000** | **0.001** | **112** | **0.000** | **0.000** | **0.008** | **5.98** | **5.93** | **0.000** | **30** | **2** | **66** |
| | | | | | | 0.002 | 0.110 | 5.40 | 5.36 | 0.000 | 38 | -8 | 56 |
| | | **0.002** | **0.082** | **29** | **0.035** | **0.004** | **0.211** | **5.21** | **5.18** | **0.000** | **14** | **-72** | **-12** |
| | | **0.002** | **0.082** | **27** | **0.041** | **0.006** | **0.238** | **5.13** | **5.10** | **0.000** | **14** | **-84** | **20** |
| | | **0.003** | **0.117** | **21** | **0.067** | **0.011** | **0.332** | **5.01** | **4.97** | **0.000** | **32** | **38** | **38** |
| | | **0.001** | **0.063** | **35** | **0.022** | **0.015** | **0.402** | **4.94** | **4.91** | **0.000** | **-12** | **-92** | **18** |
| | | | | | | 0.018 | 0.406 | 4.91 | 4.88 | 0.000 | -6 | -88 | 28 |

Height threshold: T = 4.66, p = 0.000 (0.050)
Extent threshold: k = 20 voxels, p = 0.073 (0.004)
Expected voxels per cluster, <k> = 6.285
Expected number of clusters, <c> = 0.00
FWEp: 4.661, FDRp: 5.607, FWEc: 3, FDRc: 112

Degrees of freedom = [1.0, 950.0]
FWHM = 11.3 11.3 11.6 mm mm mm; 5.6 5.7 5.8 {voxels}
Volume: 1326784 = 165848 voxels = 820.5 resels
Voxel size: 2.0 2.0 2.0 mm mm mm; (resel = 185.30 voxels)

**Table S6**

**Details of significant brain activations for Fearful > Neutral contrast in complete sample (whole-brain FWE with significance level $P_{FWE}$ < 0.05)**

| set-level | | cluster-level | | | | peak-level | | | | | mm mm mm | | |
|---|---|---|---|---|---|---|---|---|---|---|---|---|---|
| $p$ | $c$ | $p_{FWE-corr}$ | $q_{FDR-corr}$ | $k_E$ | $p_{uncorr}$ | $p_{FWE-corr}$ | $q_{FDR-corr}$ | $T$ | $(Z_E)$ | $p_{uncorr}$ | | | |
| **0.000** | **3** | **0.000** | **0.000** | **475** | **0.000** | **0.000** | **0.000** | **9.21** | **Inf** | **0.000** | **30** | **-92** | **-2** |
| | | **0.000** | **0.000** | **509** | **0.000** | **0.000** | **0.000** | **7.84** | **7.71** | **0.000** | **-34** | **-90** | **-10** |
| | | | | | | 0.000 | 0.000 | 7.31 | 7.20 | 0.000 | -28 | -98 | -4 |
| | | **0.003** | **0.067** | **21** | **0.067** | **0.001** | **0.029** | **5.44** | **5.40** | **0.000** | **50** | **32** | **6** |

Height threshold: T = 4.66, p = 0.000 (0.050)
Extent threshold: k = 0 voxels
Expected voxels per cluster, <k> = 6.285
Expected number of clusters, <c> = 0.05
FWEp: 4.661, FDRp: 5.444, FWEc: 21, FDRc: 475

Degrees of freedom = [1.0, 950.0]
FWHM = 11.3 11.3 11.6 mm mm mm; 5.6 5.7 5.8 {voxels}
Volume: 1326784 = 165848 voxels = 820.5 resels
Voxel size: 2.0 2.0 2.0 mm mm mm; (resel = 185.30 voxels)

**Table S7**

**Details of increased brain activations in LMDD compared to controls for Neutral > Baseline contrast *without* correction for antidepressant medication (cluster-level FWE with significance level $P_{FWE}$ < 0.05)**

| set-level | | cluster-level | | | | peak-level | | | | | mm mm mm | | |
|---|---|---|---|---|---|---|---|---|---|---|---|---|---|
| $p$ | $c$ | $p_{FWE-corr}$ | $q_{FDR-corr}$ | $k_E$ | $p_{uncorr}$ | $p_{FWE-corr}$ | $q_{FDR-corr}$ | $T$ | $(Z_E)$ | $p_{uncorr}$ | | | |
| **0.000** | **5** | **0.000** | **0.001** | **288** | **0.000** | **0.015** | **0.179** | **4.95** | **4.92** | **0.000** | **-36** | **18** | **40** |
| | | | | | | 0.022 | 0.179 | 4.87 | 4.84 | 0.000 | -28 | 8 | 42 |
| | | | | | | 0.275 | 0.549 | 4.20 | 4.18 | 0.000 | -20 | -2 | 44 |
| | | **0.002** | **0.012** | **150** | **0.001** | **0.020** | **0.179** | **4.90** | **4.87** | **0.000** | **26** | **2** | **42** |
| | | **0.018** | **0.082** | **83** | **0.011** | **0.024** | **0.179** | **4.85** | **4.82** | **0.000** | **-18** | **30** | **30** |
| | | **0.001** | **0.005** | **192** | **0.000** | **0.030** | **0.179** | **4.79** | **4.76** | **0.000** | **46** | **-16** | **34** |
| | | | | | | 0.239 | 0.504 | 4.25 | 4.23 | 0.000 | 58 | -8 | 26 |
| | | | | | | 0.413 | 0.679 | 4.06 | 4.04 | 0.000 | 42 | -16 | 44 |
| | | **0.000** | **0.002** | **244** | **0.000** | **0.050** | **0.230** | **4.67** | **4.64** | **0.000** | **-46** | **-62** | **22** |
| | | | | | | 0.283 | 0.549 | 4.19 | 4.17 | 0.000 | -56 | -58 | 20 |
| | | | | | | 0.724 | 0.878 | 3.80 | 3.79 | 0.000 | -50 | -68 | 34 |

Height threshold: T = 3.73, p = 0.000 (0.797)
Extent threshold: k = 80 voxels, p = 0.013 (0.020)
Expected voxels per cluster, <k> = 11.674
Expected number of clusters, <c> = 0.02
FWEp: 4.673, FDRp: Inf, FWEc: 83, FDRc: 150

Degrees of freedom = [1.0, 949.0]
FWHM = 11.1 11.1 11.4 mm mm mm; 5.5 5.6 5.7 {voxels}
Volume: 1326784 = 165848 voxels = 865.3 resels
Voxel size: 2.0 2.0 2.0 mm mm mm; (resel = 175.72 voxels)

**Table S8**

**Details of increased brain activations in LMDD compared to controls for Neutral > Baseline contrast *with* correction for antidepressant medication (cluster-level FWE with significance level $P_{FWE}$ < 0.05)**

| set-level | | cluster-level | | | | peak-level | | | | | mm mm mm | | |
|---|---|---|---|---|---|---|---|---|---|---|---|---|---|
| $p$ | $c$ | $p_{FWE-corr}$ | $q_{FDR-corr}$ | $k_E$ | $p_{uncorr}$ | $p_{FWE-corr}$ | $q_{FDR-corr}$ | $T$ | $(Z_E)$ | $p_{uncorr}$ | | | |
| **0.000** | **6** | **0.000** | **0.000** | **338** | **0.000** | **0.003** | **0.093** | **5.30** | **5.26** | **0.000** | **-28** | **10** | **42** |
| | | | | | | 0.043 | 0.273 | 4.71 | 4.68 | 0.000 | -22 | 26 | 30 |
| | | | | | | 0.064 | 0.273 | 4.61 | 4.59 | 0.000 | -36 | 16 | 40 |
| | | **0.008** | **0.062** | **106** | **0.005** | **0.019** | **0.264** | **4.90** | **4.87** | **0.000** | **38** | **-6** | **22** |
| | | | | | | 0.284 | 0.610 | 4.19 | 4.17 | 0.000 | 26 | -8 | 22 |
| | | **0.015** | **0.084** | **89** | **0.009** | **0.050** | **0.273** | **4.67** | **4.65** | **0.000** | **28** | **0** | **44** |
| | | **0.000** | **0.004** | **211** | **0.000** | **0.066** | **0.273** | **4.60** | **4.58** | **0.000** | **-48** | **-62** | **24** |
| | | | | | | 0.340 | 0.610 | 4.13 | 4.11 | 0.000 | -58 | -58 | 20 |
| | | **0.043** | **0.171** | **60** | **0.027** | **0.071** | **0.273** | **4.58** | **4.56** | **0.000** | **46** | **12** | **14** |
| | | | | | | 0.652 | 0.903 | 3.86 | 3.85 | 0.000 | 40 | 4 | 18 |
| | | **0.044** | **0.171** | **59** | **0.028** | **0.089** | **0.274** | **4.53** | **4.50** | **0.000** | **-28** | **-10** | **20** |

Height threshold: T = 3.73, p = 0.000 (0.797)
Extent threshold: k = 50 voxels, p = 0.041 (0.064)
Expected voxels per cluster, <k> = 11.687
Expected number of clusters, <c> = 0.07
FWEp: 4.673, FDRp: Inf, FWEc: 59, FDRc: 211

Degrees of freedom = [1.0, 948.0]
FWHM = 11.1 11.1 11.4 mm mm mm; 5.5 5.6 5.7 {voxels}
Volume: 1326784 = 165848 voxels = 864.3 resels
Voxel size: 2.0 2.0 2.0 mm mm mm; (resel = 175.92 voxels)

**Table S9**

**Details of increased activations in LMDD compared to controls at Fearful > Baseline contrast *without* correction for antidepressant medication (cluster-level FWE with significance level $P_{FWE}$ < 0.05)**

| set-level | | cluster-level | | | | peak-level | | | | | mm mm mm | | |
|---|---|---|---|---|---|---|---|---|---|---|---|---|---|
| $p$ | $c$ | $p_{FWE-corr}$ | $q_{FDR-corr}$ | $k_E$ | $p_{uncorr}$ | $p_{FWE-corr}$ | $q_{FDR-corr}$ | $T$ | $(Z_E)$ | $p_{uncorr}$ | | | |
| **0.000** | **4** | **0.000** | **0.000** | **1020** | **0.000** | **0.001** | **0.018** | **5.65** | **5.61** | **0.000** | **-32** | **-18** | **52** |
| | | | | | | 0.001 | 0.018 | 5.57 | 5.52 | 0.000 | -20 | -2 | 44 |
| | | | | | | 0.011 | 0.161 | 5.03 | 4.99 | 0.000 | -28 | 6 | 44 |
| | | **0.016** | **0.112** | **87** | **0.010** | **0.051** | **0.387** | **4.67** | **4.64** | **0.000** | **2** | **-16** | **44** |
| | | **0.000** | **0.004** | **225** | **0.000** | **0.062** | **0.387** | **4.62** | **4.59** | **0.000** | **-6** | **-30** | **56** |
| | | | | | | 0.124 | 0.387 | 4.44 | 4.41 | 0.000 | -4 | -28 | 64 |
| | | | | | | 0.322 | 0.653 | 4.15 | 4.13 | 0.000 | 4 | -24 | 62 |
| | | **0.002** | **0.020** | **149** | **0.001** | **0.114** | **0.387** | **4.46** | **4.44** | **0.000** | **22** | **-8** | **42** |
| | | | | | | 0.120 | 0.387 | 4.45 | 4.42 | 0.000 | 26 | 2 | 42 |

Height threshold: T = 3.73, p = 0.000 (0.797)
Extent threshold: k = 60 voxels, p = 0.027 (0.043)
Expected voxels per cluster, <k> = 11.681
Expected number of clusters, <c> = 0.04
FWEp: 4.673, FDRp: 5.571, FWEc: 87, FDRc: 149

Degrees of freedom = [1.0, 949.0]
FWHM = 11.1 11.1 11.4 mm mm mm; 5.6 5.6 5.7 {voxels}
Volume: 1326784 = 165848 voxels = 864.8 resels
Voxel size: 2.0 2.0 2.0 mm mm mm; (resel = 175.82 voxels)

**Table S10**

**Details of increased activations in LMDD compared to controls at Fearful > Baseline contrast *with* correction for antidepressant medication (cluster-level FWE with significance level $P_{FWE}$ < 0.05)**

| set-level | | cluster-level | | | | peak-level | | | | | mm mm mm | | |
|---|---|---|---|---|---|---|---|---|---|---|---|---|---|
| $p$ | $c$ | $p_{FWE-corr}$ | $q_{FDR-corr}$ | $k_E$ | $p_{uncorr}$ | $p_{FWE-corr}$ | $q_{FDR-corr}$ | $T$ | $(Z_E)$ | $p_{uncorr}$ | | | |
| **0.000** | **3** | **0.000** | **0.000** | **658** | **0.000** | **0.006** | **0.182** | **5.15** | **5.12** | **0.000** | **-20** | **-2** | **46** |
| | | | | | | 0.021 | 0.234 | 4.88 | 4.85 | 0.000 | -32 | -18 | 52 |
| | | | | | | 0.033 | 0.249 | 4.77 | 4.74 | 0.000 | -28 | 6 | 44 |
| | | **0.029** | **0.186** | **70** | **0.019** | **0.060** | **0.304** | **4.63** | **4.60** | **0.000** | **-44** | **2** | **10** |
| | | | | | | 0.235 | 0.718 | 4.25 | 4.23 | 0.000 | -40 | 8 | 14 |
| | | | | | | 0.271 | 0.718 | 4.21 | 4.19 | 0.000 | -48 | 10 | 6 |
| | | **0.020** | **0.186** | **80** | **0.013** | **0.094** | **0.360** | **4.51** | **4.49** | **0.000** | **44** | **14** | **12** |
| | | | | | | 0.430 | 0.874 | 4.05 | 4.03 | 0.000 | 48 | -2 | 10 |
| | | | | | | 0.599 | 0.925 | 3.90 | 3.89 | 0.000 | 44 | 4 | 14 |

Height threshold: T = 3.73, p = 0.000 (0.796)
Extent threshold: k = 60 voxels, p = 0.027 (0.043)
Expected voxels per cluster, <k> = 11.697
Expected number of clusters, <c> = 0.04
FWEp: 4.672, FDRp: Inf, FWEc: 70, FDRc: 658

Degrees of freedom = [1.0, 948.0]
FWHM = 11.1 11.1 11.4 mm mm mm; 5.6 5.6 5.7 {voxels}
Volume: 1326784 = 165848 voxels = 863.6 resels
Voxel size: 2.0 2.0 2.0 mm mm mm; (resel = 176.06 voxels)

**Table S11**

**Details of increased activations in LMDD compared to controls at *Neutral* > Baseline contrast with DLPFC SVC *without* correction for antidepressant medication**

| set-level | | cluster-level | | | | peak-level | | | | | mm mm mm |
|---|---|---|---|---|---|---|---|---|---|---|---|
| $p$ | $c$ | $p_{FWE\text{-}corr}$ | $q_{FDR\text{-}corr}$ | $k_E$ | $p_{uncorr}$ | $p_{FWE\text{-}corr}$ | $q_{FDR\text{-}corr}$ | $T$ | $(Z_E)$ | $p_{uncorr}$ | |
| 0.000 | 5 | 0.001 | 0.029 | 44 | 0.025 | 0.002 | 0.171 | 4.95 | 4.92 | 0.000 | -36 18 40 |
| | | | | | | 0.005 | 0.203 | 4.78 | 4.75 | 0.000 | -30 12 42 |
| | | 0.003 | 1.000 | 31 | 0.053 | 0.004 | 0.203 | 4.85 | 4.82 | 0.000 | -18 30 30 |
| | | 0.037 | 1.000 | 1 | 0.743 | 0.024 | 0.551 | 4.39 | 4.36 | 0.000 | 46 6 14 |
| | | 0.027 | 1.000 | 3 | 0.539 | 0.028 | 1.000 | 4.35 | 4.33 | 0.000 | 46 12 16 |
| | | 0.021 | 1.000 | 5 | 0.419 | 0.029 | 1.000 | 4.35 | 4.32 | 0.000 | 40 12 36 |

Height threshold: T = 4.20, p = 0.000 (0.050)
Extent threshold: k = 0 voxels
Expected voxels per cluster, <k> = 8.193
Expected number of clusters, <c> = 0.05
FWEp: 4.195, FDRp: Inf, FWEc: 41, FDRc: 41

Degrees of freedom = [1.0, 949.0]
FWHM = 11.1 11.1 11.4 mm mm mm; 5.5 5.6 5.7 {voxels}
Volume: 130992 = 16374 voxels = 105.2 resels
Voxel size: 2.0 2.0 2.0 mm mm mm; (resel = 175.72 voxels)

**Table S12**

**Details of increased activations in LMDD compared to controls at *Neutral* > Baseline contrast with DLPFC SVC *with* correction for antidepressant medication**

| set-level | | cluster-level | | | | peak-level | | | | | mm mm mm |
|---|---|---|---|---|---|---|---|---|---|---|---|
| $p$ | $c$ | $p_{FWE\text{-}corr}$ | $q_{FDR\text{-}corr}$ | $k_E$ | $p_{uncorr}$ | $p_{FWE\text{-}corr}$ | $q_{FDR\text{-}corr}$ | $T$ | $(Z_E)$ | $p_{uncorr}$ | |
| 0.000 | 4 | 0.006 | 1.000 | 20 | 0.112 | 0.002 | 0.293 | 4.98 | 4.95 | 0.000 | -30 12 42 |
| | | 0.004 | 0.090 | 24 | 0.084 | 0.007 | 0.293 | 4.71 | 4.68 | 0.000 | -22 26 30 |
| | | 0.021 | 1.000 | 5 | 0.419 | 0.014 | 0.293 | 4.52 | 4.50 | 0.000 | 46 10 14 |
| | | 0.031 | 1.000 | 2 | 0.624 | 0.021 | 1.000 | 4.43 | 4.40 | 0.000 | 46 6 14 |

Height threshold: T = 4.19, p = 0.000 (0.050)
Extent threshold: k = 0 voxels
Expected voxels per cluster, <k> = 8.203
Expected number of clusters, <c> = 0.05
FWEp: 4.195, FDRp: Inf, FWEc: 23, FDRc: Inf

Degrees of freedom = [1.0, 948.0]
FWHM = 11.1 11.1 11.4 mm mm mm; 5.5 5.6 5.7 {voxels}
Volume: 130992 = 16374 voxels = 105.1 resels
Voxel size: 2.0 2.0 2.0 mm mm mm; (resel = 175.92 voxels)

**Table S13**

**Details of increased activations in LMDD compared to controls at *Fearful* > Baseline contrast with DLPFC SVC *without* correction for antidepressant medication**

| cluster-level | | | | peak-level | | | | | mm mm mm |
|---|---|---|---|---|---|---|---|---|---|
| $p_{FWE-corr}$ | $q_{FDR-corr}$ | $k_E$ | $p_{uncorr}$ | $p_{FWE-corr}$ | $q_{FDR-corr}$ | $T$ | $(Z_E)$ | $p_{uncorr}$ | |
| **0.002** | **0.151** | **33** | **0.047** | **0.003** | **0.175** | **4.92** | **4.89** | **0.000** | **-30 8 42** |
| | | | | 0.010 | 0.292 | 4.61 | 4.58 | 0.000 | -36 16 38 |

Height threshold: T = 4.20, p = 0.000 (0.050)
Extent threshold: k = 0 voxels
Expected voxels per cluster, <k> = 8.198
Expected number of clusters, <c> = 0.05
FWEp: 4.195, FDRp: Inf, FWEc: 16, FDRc: Inf

Degrees of freedom = [1.0, 949.0]
FWHM = 11.1 11.1 11.4 mm mm mm; 5.6 5.6 5.7 {voxels}
Volume: 130992 = 16374 voxels = 105.1 resels
Voxel size: 2.0 2.0 2.0 mm mm mm; (resel = 175.82 voxels)

**Table S14**

**Details of increased activations in LMDD compared to controls at *Fearful* > Baseline contrast with DLPFC SVC *with* correction for antidepressant medication**

| cluster-level | | | | peak-level | | | | | mm mm mm |
|---|---|---|---|---|---|---|---|---|---|
| $p_{FWE-corr}$ | $q_{FDR-corr}$ | $k_E$ | $p_{uncorr}$ | $p_{FWE-corr}$ | $q_{FDR-corr}$ | $T$ | $(Z_E)$ | $p_{uncorr}$ | |
| **0.017** | **0.743** | **7** | **0.337** | **0.011** | **0.405** | **4.60** | **4.57** | **0.000** | **-30 10 42** |

Height threshold: T = 4.19, p = 0.000 (0.050)
Extent threshold: k = 0 voxels
Expected voxels per cluster, <k> = 8.212
Expected number of clusters, <c> = 0.05
FWEp: 4.195, FDRp: Inf, FWEc: 1, FDRc: Inf

Degrees of freedom = [1.0, 948.0]
FWHM = 11.1 11.1 11.4 mm mm mm; 5.6 5.6 5.7 {voxels}
Volume: 130992 = 16374 voxels = 105.0 resels
Voxel size: 2.0 2.0 2.0 mm mm mm; (resel = 176.06 voxels)

**Table S15**

**Estimates and estimate probabilities of parameters of full DCM model (both endogenous connections and modulatory inputs), with LMDD as the main predictor of interest and *without* medication covariate**

| Model Parameter | Estimate Common | Estimate Age | Estimate Sex | Estimate Site | Estimate LMDD | Probability Common | Probability Age | Probability Sex | Probability Site | Probability LMDD |
|---|---|---|---|---|---|---|---|---|---|---|
| Base \| V1 → V1 | 0.205 | 0 | 0 | 0 | 0 | 1 | 0 | 0 | 0 | 0 |
| Base \| V1 → Amyg | 0.151 | 0 | 0 | 0.071 | 0 | 1 | 0 | 0 | 1 | 0 |
| Base \| V1 → DLPFC | 0.169 | 0 | 0 | 0 | 0 | 1 | 0 | 0 | 0 | 0 |
| Base \| Amyg → V1 | -0.479 | 0 | 0.125 | 0 | 0 | 1 | 0 | 0.821 | 0 | 0 |
| Base \| Amyg → Amyg | -0.617 | 0 | 0 | 0.143 | 0 | 1 | 0 | 0 | 0.991 | 0 |
| Base \| Amyg → DLPFC | -0.183 | 0 | 0 | 0 | 0 | 1 | 0 | 0 | 0 | 0 |
| Base \| DLPFC → V1 | -0.297 | 0 | 0 | 0.327 | 0 | 1 | 0 | 0 | 1 | 0 |
| Base \| DLPFC → Amyg | -0.053 | 0 | 0 | 0 | 0 | 1 | 0 | 0 | 0 | 0 |
| Base \| DLPFC → DLPFC | -0.521 | 0 | 0.082 | 0 | 0 | 1 | 0 | 0.797 | 0 | 0 |
| Neutral modulation \| V1 → V1 | 0 | 0 | 0 | 0 | 0 | 0 | 0 | 0 | 0 | 0 |
| Neutral modulation \| V1 → Amyg | 0 | 0 | 0 | 0 | 0 | 0 | 0 | 0 | 0 | 0 |
| Neutral modulation \| V1 → DLPFC | -0.128 | 0 | 0 | 0 | 0 | 1 | 0 | 0 | 0 | 0 |
| Neutral modulation \| Amyg → V1 | 0.77 | 0 | 0 | 0.473 | 0 | 1 | 0 | 0 | 0.899 | 0 |
| Neutral modulation \| Amyg → Amyg | 0 | 0 | 0 | 0 | 0 | 0 | 0 | 0 | 0 | 0 |
| Neutral modulation \| Amyg → DLPFC | 0.109 | 0 | 0 | 0 | 0 | 1 | 0 | 0 | 0 | 0 |
| Neutral modulation \| DLPFC → V1 | 0.361 | 0 | 0 | 0 | 0 | 1 | 0 | 0 | 0 | 0 |
| Neutral modulation \| DLPFC → Amyg | 0 | 0 | 0 | 0 | 0 | 0 | 0 | 0 | 0 | 0 |
| Neutral modulation \| DLPFC → DLPFC | 0 | 0 | 0 | 0 | 0 | 0 | 0 | 0 | 0 | 0 |
| Fearful modulation \| V1 → V1 | -0.446 | 0 | 0 | 0.216 | 0 | 1 | 0 | 0 | 0.634 | 0 |
| Fearful modulation \| V1 → Amyg | 0 | 0 | 0 | 0 | 0 | 0 | 0 | 0 | 0 | 0 |
| Fearful modulation \| V1 → DLPFC | -0.126 | 0 | 0 | 0 | 0.065 | 1 | 0 | 0 | 0 | 0.579 |
| Fearful modulation \| Amyg → V1 | 0.432 | 0 | 0 | 0 | 0 | 1 | 0 | 0 | 0 | 0 |
| Fearful modulation \| Amyg → Amyg | 0.25 | 0 | 0 | 0 | 0 | 1 | 0 | 0 | 0 | 0 |
| Fearful modulation \| Amyg → DLPFC | 0 | 0 | 0 | 0 | -0.151 | 0 | 0 | 0 | 0 | 0.818 |
| Fearful modulation \| DLPFC → V1 | 0.371 | 0 | 0 | 0 | 0 | 1 | 0 | 0 | 0 | 0 |
| Fearful modulation \| DLPFC → Amyg | 0 | 0 | 0 | 0 | 0 | 0 | 0 | 0 | 0 | 0 |
| Fearful modulation \| DLPFC → DLPFC | 0.245 | 0 | 0 | 0 | 0 | 1 | 0 | 0 | 0 | 0 |

***Note***: Rows with non-zero probability associations of LMDD highlighted in light blue.

**Table S16**

**Estimates and estimate probabilities of parameters of full DCM model (both endogenous connections and modulatory inputs), with LMDD as the main predictor of interest and *with* medication covariate**

| Model Parameter | Estimate Common | Estimate Age | Estimate Sex | Estimate Site | Estimate Medicated | Estimate LMDD | Probability Common | Probability Age | Probability Sex | Probability Site | Probability Medicated | Probability LMDD |
|---|---|---|---|---|---|---|---|---|---|---|---|---|
| Base \| V1 → V1 | 0.2 | 0 | 0 | 0 | 0 | 0 | 1 | 0 | 0 | 0 | 0 | 0 |
| Base \| V1 → Amyg | 0.151 | 0 | 0 | 0.071 | 0 | 0 | 1 | 0 | 0 | 1 | 0 | 0 |
| Base \| V1 → DLPFC | 0.168 | 0 | 0 | 0 | 0 | 0 | 1 | 0 | 0 | 0 | 0 | 0 |
| Base \| Amyg → V1 | -0.479 | 0 | 0.125 | 0 | 0 | 0 | 1 | 0 | 0.82 | 0 | 0 | 0 |
| Base \| Amyg → Amyg | -0.618 | 0 | 0 | 0.145 | 0 | 0 | 1 | 0 | 0 | 0.991 | 0 | 0 |
| Base \| Amyg → DLPFC | -0.183 | 0 | 0 | 0 | 0 | 0 | 1 | 0 | 0 | 0 | 0 | 0 |
| Base \| DLPFC → V1 | -0.296 | 0 | 0 | 0.324 | 0 | 0 | 1 | 0 | 0 | 1 | 0 | 0 |
| Base \| DLPFC → Amyg | -0.053 | 0 | 0 | 0 | 0 | 0 | 1 | 0 | 0 | 0 | 0 | 0 |
| Base \| DLPFC → DLPFC | -0.523 | 0 | 0.082 | 0 | 0 | 0 | 1 | 0 | 0.797 | 0 | 0 | 0 |
| Neutral modulation \| V1 → V1 | 0 | 0 | 0 | 0 | 0 | 0 | 0 | 0 | 0 | 0 | 0 | 0 |
| Neutral modulation \| V1 → Amyg | 0 | 0 | 0 | 0 | 0 | 0 | 0 | 0 | 0 | 0 | 0 | 0 |
| Neutral modulation \| V1 → DLPFC | -0.127 | 0 | 0 | 0 | 0 | 0 | 1 | 0 | 0 | 0 | 0 | 0 |
| Neutral modulation \| Amyg → V1 | 0.764 | 0 | 0 | 0.473 | 0 | 0 | 1 | 0 | 0 | 0.9 | 0 | 0 |
| Neutral modulation \| Amyg → Amyg | 0 | 0 | 0 | 0 | 0 | 0 | 0 | 0 | 0 | 0 | 0 | 0 |
| Neutral modulation \| Amyg → DLPFC | 0.109 | 0 | 0 | 0 | 0 | 0 | 1 | 0 | 0 | 0 | 0 | 0 |
| Neutral modulation \| DLPFC → V1 | 0.361 | 0 | 0 | 0 | 0 | 0 | 1 | 0 | 0 | 0 | 0 | 0 |
| Neutral modulation \| DLPFC → Amyg | 0 | 0 | 0 | 0 | 0 | 0 | 0 | 0 | 0 | 0 | 0 | 0 |
| Neutral modulation \| DLPFC → DLPFC | 0 | 0 | 0 | 0 | 0 | 0 | 0 | 0 | 0 | 0 | 0 | 0 |
| Fearful modulation \| V1 → V1 | -0.444 | 0 | 0 | 0.222 | 0 | 0 | 1 | 0 | 0 | 0.634 | 0 | 0 |
| Fearful modulation \| V1 → Amyg | 0 | 0 | 0 | 0 | 0 | 0 | 0 | 0 | 0 | 0 | 0 | 0 |
| Fearful modulation \| V1 → DLPFC | -0.128 | 0 | 0 | 0 | 0 | 0.056 | 1 | 0 | 0 | 0 | 0 | 0.578 |
| Fearful modulation \| Amyg → V1 | 0.429 | 0 | 0 | 0 | 0 | 0 | 1 | 0 | 0 | 0 | 0 | 0 |
| Fearful modulation \| Amyg → Amyg | 0.252 | 0 | 0 | 0 | 0 | 0 | 1 | 0 | 0 | 0 | 0 | 0 |
| Fearful modulation \| Amyg → DLPFC | 0 | 0 | 0 | 0 | 0 | -0.149 | 0 | 0 | 0 | 0 | 0 | 0.818 |
| Fearful modulation \| DLPFC → V1 | 0.372 | 0 | 0 | 0 | 0 | 0 | 1 | 0 | 0 | 0 | 0 | 0 |
| Fearful modulation \| DLPFC → Amyg | 0 | 0 | 0 | 0 | 0 | 0 | 0 | 0 | 0 | 0 | 0 | 0 |
| Fearful modulation \| DLPFC → DLPFC | 0.246 | 0 | 0 | 0 | 0 | 0 | 1 | 0 | 0 | 0 | 0 | 0 |

***Note***: Rows with non-zero probability associations of LMDD highlighted in light blue. No associations of antidepressant medication were observed.

**Table S17**

**Estimates and estimate probabilities of parameters of DCM model with only endogenous connections, with LMDD as the main predictor of interest and *without* medication covariate**

| Model Parameter | Estimate Common | Estimate Age | Estimate Sex | Estimate Site | Estimate LMDD | Probability Common | Probability Age | Probability Sex | Probability Site | Probability LMDD |
|---|---|---|---|---|---|---|---|---|---|---|
| Base \| V1 → V1 | 0.18 | -0.009 | 0 | 0 | 0 | 1 | 0.978 | 0 | 0 | 0 |
| Base \| V1 → Amyg | 0.183 | 0.003 | 0 | 0.091 | 0 | 1 | 0.899 | 0 | 0.996 | 0 |
| Base \| V1 → DLPFC | 0.211 | 0 | 0 | 0 | 0 | 1 | 0 | 0 | 0 | 0 |
| Base \| Amyg → V1 | -0.485 | 0 | 0.112 | 0 | 0 | 1 | 0 | 0.802 | 0 | 0 |
| Base \| Amyg → Amyg | -0.475 | 0 | 0 | 0.188 | 0 | 1 | 0 | 0 | 1 | 0 |
| Base \| Amyg → DLPFC | -0.218 | 0 | 0 | 0 | 0 | 1 | 0 | 0 | 0 | 0 |
| Base \| DLPFC → V1 | -0.305 | 0 | 0 | 0.341 | 0 | 1 | 0 | 0 | 1 | 0 |
| Base \| DLPFC → Amyg | -0.07 | 0 | 0 | 0 | 0 | 1 | 0 | 0 | 0 | 0 |
| Base \| DLPFC → DLPFC | -0.373 | 0 | 0.141 | 0 | 0 | 1 | 0 | 0.999 | 0 | 0 |

***Note***: No associations of LMDD were observed.

**Table S18**

**Estimates and estimate probabilities of parameters of DCM model with only endogenous connections, with LMDD as the main predictor of interest and *with* medication covariate**

| Model Parameter | Estimate Common | Estimate Age | Estimate Sex | Estimate Site | Estimate Medicated | Estimate LMDD | Probability Common | Probability Age | Probability Sex | Probability Site | Probability Medicated | Probability LMDD |
|---|---|---|---|---|---|---|---|---|---|---|---|---|
| Base \| V1 → V1 | 0.18 | -0.009 | 0 | 0 | 0 | 0 | 1 | 0.979 | 0 | 0 | 0 | 0 |
| Base \| V1 → Amyg | 0.183 | 0.003 | 0 | 0.092 | 0 | 0 | 1 | 0.905 | 0 | 0.996 | 0 | 0 |
| Base \| V1 → DLPFC | 0.212 | 0 | 0 | 0 | 0 | 0 | 1 | 0 | 0 | 0 | 0 | 0 |
| Base \| Amyg → V1 | -0.484 | 0 | 0.117 | 0 | 0 | 0 | 1 | 0 | 0.806 | 0 | 0 | 0 |
| Base \| Amyg → Amyg | -0.474 | 0 | 0 | 0.185 | 0 | 0 | 1 | 0 | 0 | 0.999 | 0 | 0 |
| Base \| Amyg → DLPFC | -0.216 | 0 | 0 | 0 | 0 | 0 | 1 | 0 | 0 | 0 | 0 | 0 |
| Base \| DLPFC → V1 | -0.306 | 0 | 0 | 0.339 | 0 | 0 | 1 | 0 | 0 | 1 | 0 | 0 |
| Base \| DLPFC → Amyg | -0.07 | 0 | 0 | 0 | 0 | 0 | 1 | 0 | 0 | 0 | 0 | 0 |
| Base \| DLPFC → DLPFC | -0.369 | 0 | 0.119 | 0 | 0 | 0 | 1 | 0 | 0.941 | 0 | 0 | 0 |

***Note***: No associations of LMDD or of antidepressant medication status were observed.

**Table S19**

**Estimates and estimate probabilities of parameters of DCM model with only modulatory inputs, with LMDD as the main predictor of interest and *without* medication covariate**

| Model Parameter | Estimate Common | Estimate Age | Estimate Sex | Estimate Site | Estimate LMDD | Probability Common | Probability Age | Probability Sex | Probability Site | Probability LMDD |
|---|---|---|---|---|---|---|---|---|---|---|
| Neutral modulation \| V1 → V1 | 0.3 | 0 | 0 | 0 | 0 | 1 | 0 | 0 | 0 | 0 |
| Neutral modulation \| V1 → Amyg | 0 | 0 | 0 | 0 | 0 | 0 | 0 | 0 | 0 | 0 |
| Neutral modulation \| V1 → DLPFC | -0.258 | 0 | 0 | 0 | 0 | 1 | 0 | 0 | 0 | 0 |
| Neutral modulation \| Amyg → V1 | 1.044 | 0 | 0 | 0.182 | 0 | 1 | 0 | 0 | 0.526 | 0 |
| Neutral modulation \| Amyg → Amyg | 0.345 | 0 | 0 | 0 | 0 | 1 | 0 | 0 | 0 | 0 |
| Neutral modulation \| Amyg → DLPFC | 0.149 | 0 | 0 | 0 | 0 | 1 | 0 | 0 | 0 | 0 |
| Neutral modulation \| DLPFC → V1 | 0.624 | 0 | 0 | -0.381 | 0 | 1 | 0 | 0 | 0.885 | 0 |
| Neutral modulation \| DLPFC → Amyg | 0 | 0 | 0 | 0 | 0 | 0 | 0 | 0 | 0 | 0 |
| Neutral modulation \| DLPFC → DLPFC | 0.11 | 0 | 0 | 0 | 0 | 0.797 | 0 | 0 | 0 | 0 |
| Fearful modulation \| V1 → V1 | 0 | 0 | 0 | 0 | 0 | 0 | 0 | 0 | 0 | 0 |
| Fearful modulation \| V1 → Amyg | 0 | 0 | 0 | 0 | 0 | 0 | 0 | 0 | 0 | 0 |
| Fearful modulation \| V1 → DLPFC | -0.298 | 0 | -0.088 | 0 | 0 | 1 | 0 | 0.619 | 0 | 0 |
| Fearful modulation \| Amyg → V1 | 0.562 | 0 | 0 | 0 | 0 | 1 | 0 | 0 | 0 | 0 |
| Fearful modulation \| Amyg → Amyg | 0.49 | 0 | 0 | 0 | 0 | 1 | 0 | 0 | 0 | 0 |
| Fearful modulation \| Amyg → DLPFC | 0 | 0 | 0 | 0 | -0.131 | 0 | 0 | 0 | 0 | 0.614 |
| Fearful modulation \| DLPFC → V1 | 0.651 | 0 | 0 | 0 | 0 | 1 | 0 | 0 | 0 | 0 |
| Fearful modulation \| DLPFC → Amyg | 0 | 0 | 0 | 0 | 0 | 0 | 0 | 0 | 0 | 0 |
| Fearful modulation \| DLPFC → DLPFC | 0.393 | 0 | 0 | 0 | 0 | 1 | 0 | 0 | 0 | 0 |

***Note***: Row with non-zero probability association of LMDD highlighted in light blue.

**Table S20**

**Estimates and estimate probabilities of parameters of DCM model with only modulatory inputs, with LMDD as the main predictor of interest and *with* medication covariate**

| Model Parameter | Estimate Common | Estimate Age | Estimate Sex | Estimate Site | Estimate Medicated | Estimate LMDD | Probability Common | Probability Age | Probability Sex | Probability Site | Probability Medicated | Probability LMDD |
|---|---|---|---|---|---|---|---|---|---|---|---|---|
| Neutral modulation \| V1 → V1 | 0.3 | 0 | 0 | 0 | 0 | 0 | 1 | 0 | 0 | 0 | 0 | 0 |
| Neutral modulation \| V1 → Amyg | 0 | 0 | 0 | 0 | 0 | 0 | 0 | 0 | 0 | 0 | 0 | 0 |
| Neutral modulation \| V1 → DLPFC | -0.258 | 0 | 0 | 0 | 0 | 0 | 1 | 0 | 0 | 0 | 0 | 0 |
| Neutral modulation \| Amyg → V1 | 1.045 | 0 | 0 | 0.179 | 0 | 0 | 1 | 0 | 0 | 0.527 | 0 | 0 |
| Neutral modulation \| Amyg → Amyg | 0.345 | 0 | 0 | 0 | 0 | 0 | 1 | 0 | 0 | 0 | 0 | 0 |
| Neutral modulation \| Amyg → DLPFC | 0.15 | 0 | 0 | 0 | 0 | 0 | 1 | 0 | 0 | 0 | 0 | 0 |
| Neutral modulation \| DLPFC → V1 | 0.624 | 0 | 0 | -0.38 | 0 | 0 | 1 | 0 | 0 | 0.886 | 0 | 0 |
| Neutral modulation \| DLPFC → Amyg | 0 | 0 | 0 | 0 | 0 | 0 | 0 | 0 | 0 | 0 | 0 | 0 |
| Neutral modulation \| DLPFC → DLPFC | 0.107 | 0 | 0 | 0 | 0 | 0 | 0.799 | 0 | 0 | 0 | 0 | 0 |
| Fearful modulation \| V1 → V1 | 0 | 0 | 0 | 0 | 0 | 0 | 0 | 0 | 0 | 0 | 0 | 0 |
| Fearful modulation \| V1 → Amyg | 0 | 0 | 0 | 0 | 0 | 0 | 0 | 0 | 0 | 0 | 0 | 0 |
| Fearful modulation \| V1 → DLPFC | -0.299 | 0 | -0.089 | 0 | 0 | 0 | 1 | 0 | 0.618 | 0 | 0 | 0 |
| Fearful modulation \| Amyg → V1 | 0.561 | 0 | 0 | 0 | 0 | 0 | 1 | 0 | 0 | 0 | 0 | 0 |
| Fearful modulation \| Amyg → Amyg | 0.495 | 0 | 0 | 0 | 0 | 0 | 1 | 0 | 0 | 0 | 0 | 0 |
| Fearful modulation \| Amyg → DLPFC | 0 | 0 | 0 | 0 | 0 | -0.122 | 0 | 0 | 0 | 0 | 0 | 0.613 |
| Fearful modulation \| DLPFC → V1 | 0.647 | 0 | 0 | 0 | 0 | 0 | 1 | 0 | 0 | 0 | 0 | 0 |
| Fearful modulation \| DLPFC → Amyg | 0 | 0 | 0 | 0 | 0 | 0 | 0 | 0 | 0 | 0 | 0 | 0 |
| Fearful modulation \| DLPFC → DLPFC | 0.395 | 0 | 0 | 0 | 0 | 0 | 1 | 0 | 0 | 0 | 0 | 0 |

***Note***: Row with non-zero probability association of LMDD highlighted in light blue. No associations of antidepressant medication were observed.

**Table S21**

**Estimates and estimate probabilities of parameters of full DCM model (both endogenous connections and modulatory inputs), with QIDS score as the main predictor of interest (no medication covariate)**

| Model Parameter | Estimate Common | Estimate Age | Estimate Sex | Estimate Site | Estimate QIDS | Probability Common | Probability Age | Probability Sex | Probability Site | Probability QIDS |
|---|---|---|---|---|---|---|---|---|---|---|
| Base \| V1 → V1 | 0.21 | 0 | 0 | 0 | 0 | 1 | 0 | 0 | 0 | 0 |
| Base \| V1 → Amyg | 0.151 | 0 | 0 | 0.072 | 0 | 1 | 0 | 0 | 0.999 | 0 |
| Base \| V1 → DLPFC | 0.168 | 0 | 0 | 0 | 0 | 1 | 0 | 0 | 0 | 0 |
| Base \| Amyg → V1 | -0.479 | 0 | 0.113 | 0 | 0 | 1 | 0 | 0.792 | 0 | 0 |
| Base \| Amyg → Amyg | -0.617 | 0 | 0 | 0.149 | 0 | 1 | 0 | 0 | 0.993 | 0 |
| Base \| Amyg → DLPFC | -0.182 | 0 | 0 | 0 | 0 | 1 | 0 | 0 | 0 | 0 |
| Base \| DLPFC → V1 | -0.294 | 0 | 0 | 0.325 | 0 | 1 | 0 | 0 | 1 | 0 |
| Base \| DLPFC → Amyg | -0.053 | 0 | 0 | 0 | 0 | 1 | 0 | 0 | 0 | 0 |
| Base \| DLPFC → DLPFC | -0.522 | 0 | 0.091 | 0 | 0 | 1 | 0 | 0.823 | 0 | 0 |
| Neutral modulation \| V1 → V1 | 0 | 0 | 0 | 0 | 0 | 0 | 0 | 0 | 0 | 0 |
| Neutral modulation \| V1 → Amyg | 0 | 0 | 0 | 0 | 0 | 0 | 0 | 0 | 0 | 0 |
| Neutral modulation \| V1 → DLPFC | -0.128 | 0 | 0 | 0 | 0 | 1 | 0 | 0 | 0 | 0 |
| Neutral modulation \| Amyg → V1 | 0.764 | 0 | 0 | 0.454 | 0 | 1 | 0 | 0 | 0.885 | 0 |
| Neutral modulation \| Amyg → Amyg | 0 | 0 | 0 | 0 | 0 | 0 | 0 | 0 | 0 | 0 |
| Neutral modulation \| Amyg → DLPFC | 0.11 | 0 | 0 | 0 | 0 | 1 | 0 | 0 | 0 | 0 |
| Neutral modulation \| DLPFC → V1 | 0.362 | 0 | 0 | 0 | 0 | 1 | 0 | 0 | 0 | 0 |
| Neutral modulation \| DLPFC → Amyg | 0 | 0 | 0 | 0 | 0 | 0 | 0 | 0 | 0 | 0 |
| Neutral modulation \| DLPFC → DLPFC | 0 | 0 | 0 | 0 | 0 | 0 | 0 | 0 | 0 | 0 |
| Fearful modulation \| V1 → V1 | -0.445 | 0 | 0 | 0.214 | 0 | 1 | 0 | 0 | 0.626 | 0 |
| Fearful modulation \| V1 → Amyg | 0 | 0 | 0 | 0 | 0 | 0 | 0 | 0 | 0 | 0 |
| Fearful modulation \| V1 → DLPFC | -0.126 | 0 | 0 | 0 | 0 | 1 | 0 | 0 | 0 | 0 |
| Fearful modulation \| Amyg → V1 | 0.429 | 0 | 0 | 0 | 0 | 1 | 0 | 0 | 0 | 0 |
| Fearful modulation \| Amyg → Amyg | 0.252 | 0 | 0 | 0 | 0 | 1 | 0 | 0 | 0 | 0 |
| Fearful modulation \| Amyg → DLPFC | 0 | 0 | 0 | 0 | 0 | 0 | 0 | 0 | 0 | 0 |
| Fearful modulation \| DLPFC → V1 | 0.369 | 0 | 0 | 0 | 0 | 1 | 0 | 0 | 0 | 0 |
| Fearful modulation \| DLPFC → Amyg | 0 | 0 | 0 | 0 | 0 | 0 | 0 | 0 | 0 | 0 |
| Fearful modulation \| DLPFC → DLPFC | 0.25 | 0 | 0 | 0 | 0 | 1 | 0 | 0 | 0 | 0 |

***Note***: No associations of QIDS scores were observed.

**Table S22**

**Estimates and estimate probabilities of parameters of DCM model with only endogenous connections, with QIDS score as the main predictor of interest (no medication covariate)**

| Model Parameter | Estimate Common | Estimate Age | Estimate Sex | Estimate Site | Estimate QIDS | Probability Common | Probability Age | Probability Sex | Probability Site | Probability QIDS |
|---|---|---|---|---|---|---|---|---|---|---|
| Base \| V1 → V1 | 0.21 | -0.01 | 0 | 0 | 0 | 1 | 1 | 0 | 0 | 0 |
| Base \| V1 → Amyg | 0.18 | 0 | 0 | 0.08 | 0 | 1 | 0.85 | 0 | 0.98 | 0 |
| Base \| V1 → DLPFC | 0.21 | 0 | 0 | 0 | 0 | 1 | 0 | 0 | 0 | 0 |
| Base \| Amyg → V1 | -0.49 | 0 | 0.09 | 0 | 0 | 1 | 0 | 0.73 | 0 | 0 |
| Base \| Amyg → Amyg | -0.47 | 0 | 0 | 0.21 | 0 | 1 | 0.86 | 0 | 1 | 0 |
| Base \| Amyg → DLPFC | -0.21 | 0 | 0 | 0 | 0 | 1 | 0 | 0 | 0 | 0 |
| Base \| DLPFC → V1 | -0.29 | 0 | 0 | 0.33 | 0 | 1 | 0 | 0 | 1 | 0 |
| Base \| DLPFC → Amyg | -0.06 | 0 | 0 | 0 | 0 | 1 | 0 | 0 | 0 | 0 |
| Base \| DLPFC → DLPFC | -0.36 | 0 | 0.16 | -0.05 | 0 | 1 | 0 | 1 | 0.76 | 0 |

***Note***: No associations of QIDS scores were observed.

**Table S23**

**Estimates and estimate probabilities of parameters of DCM model with only modulatory inputs, with QIDS score as the main predictor of interest (no medication covariate)**

| Model Parameter | Estimate Common | Estimate Age | Estimate Sex | Estimate Site | Estimate QIDS | Probability Common | Probability Age | Probability Sex | Probability Site | Probability QIDS |
|---|---|---|---|---|---|---|---|---|---|---|
| Neutral modulation \| V1 → V1 | 0.3 | 0 | 0 | 0 | 0 | 1 | 0 | 0 | 0 | 0 |
| Neutral modulation \| V1 → Amyg | 0 | 0 | 0 | 0 | 0 | 0 | 0 | 0 | 0 | 0 |
| Neutral modulation \| V1 → DLPFC | -0.257 | 0 | 0 | 0 | 0 | 1 | 0 | 0 | 0 | 0 |
| Neutral modulation \| Amyg → V1 | 1.045 | 0 | 0 | 0.177 | 0 | 1 | 0 | 0 | 0.525 | 0 |
| Neutral modulation \| Amyg → Amyg | 0.346 | 0 | 0 | 0 | 0 | 1 | 0 | 0 | 0 | 0 |
| Neutral modulation \| Amyg → DLPFC | 0.151 | 0 | 0 | 0 | 0 | 1 | 0 | 0 | 0 | 0 |
| Neutral modulation \| DLPFC → V1 | 0.624 | 0 | 0 | -0.369 | 0 | 1 | 0 | 0 | 0.885 | 0 |
| Neutral modulation \| DLPFC → Amyg | 0 | 0 | 0 | 0 | 0 | 0 | 0 | 0 | 0 | 0 |
| Neutral modulation \| DLPFC → DLPFC | 0.112 | 0 | 0 | 0 | 0.02 | 0.793 | 0 | 0 | 0 | 0.606 |
| Fearful modulation \| V1 → V1 | 0 | 0 | 0 | 0 | 0 | 0 | 0 | 0 | 0 | 0 |
| Fearful modulation \| V1 → Amyg | 0 | 0 | 0 | 0 | 0.012 | 0 | 0 | 0 | 0 | 0.645 |
| Fearful modulation \| V1 → DLPFC | -0.298 | 0 | -0.082 | 0 | 0 | 1 | 0 | 0.611 | 0 | 0 |
| Fearful modulation \| Amyg → V1 | 0.56 | 0 | 0 | 0 | 0 | 1 | 0 | 0 | 0 | 0 |
| Fearful modulation \| Amyg → Amyg | 0.492 | 0 | 0 | 0 | 0 | 1 | 0 | 0 | 0 | 0 |
| Fearful modulation \| Amyg → DLPFC | 0 | 0 | 0 | 0 | 0 | 0 | 0 | 0 | 0 | 0 |
| Fearful modulation \| DLPFC → V1 | 0.649 | 0 | 0 | 0 | 0 | 1 | 0 | 0 | 0 | 0 |
| Fearful modulation \| DLPFC → Amyg | 0 | 0 | 0 | 0 | 0 | 0 | 0 | 0 | 0 | 0 |
| Fearful modulation \| DLPFC → DLPFC | 0.393 | 0 | 0 | 0 | 0 | 1 | 0 | 0 | 0 | 0 |

***Note***: Rows with non-zero probability associations of QIDS scores highlighted in light blue.

**Figure S1 Amygdala region-of-interest.** Amygdala ROI applied in SVC analyses and in analyses of mean amygdala activation – overlaid over second-level mask (A) and over average brain template (B).

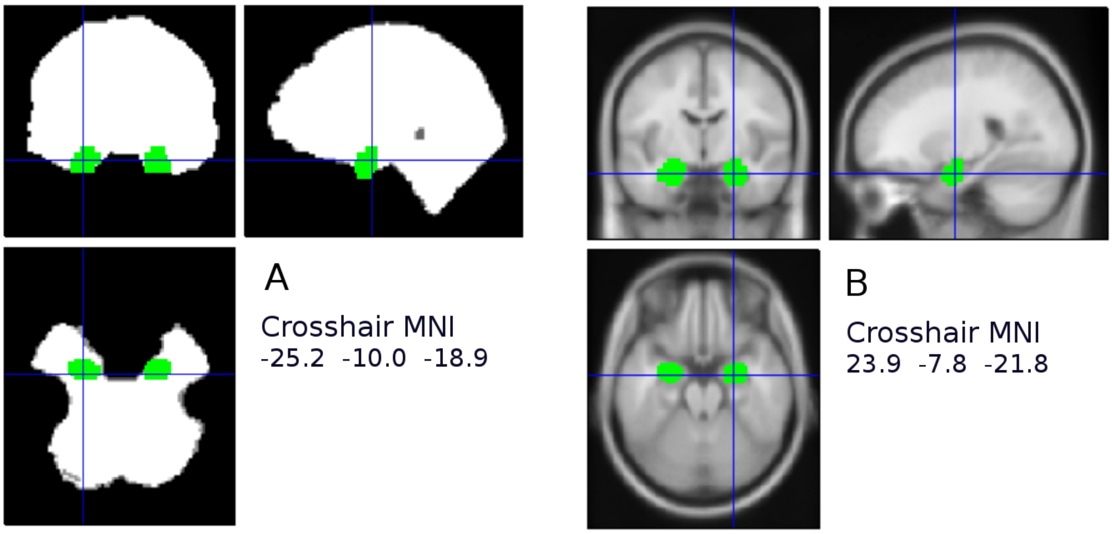


**Figure S2 Amygdala signal dropout.** Signal dropout in the caudal part of the amygdala ROI. Second-level mask is presented blue, amygdala ROI without signal coverage in white.

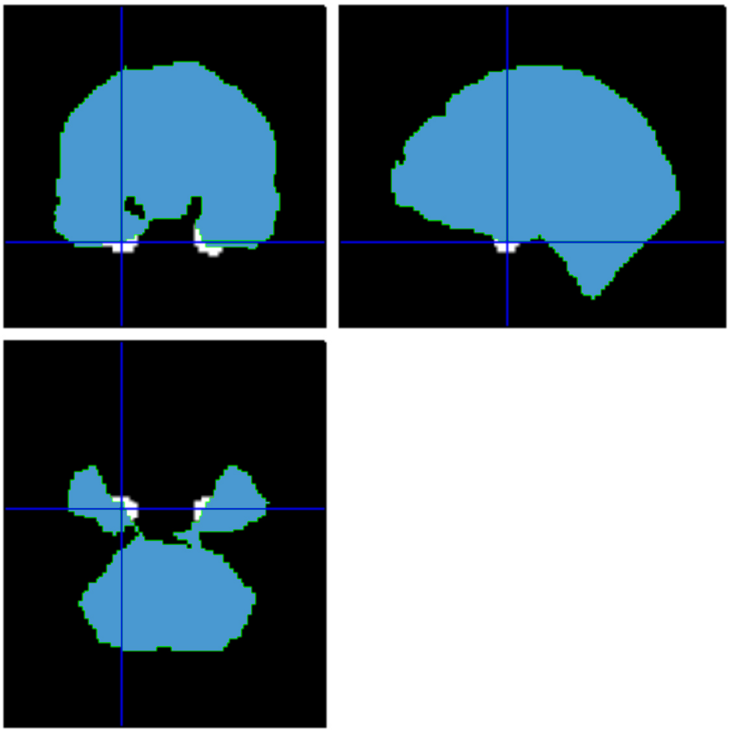

**Figure S3 Brain activation in response to face stimuli.** 'Glass' brain illustration of activations in the complete sample ( $N = 954$ ) in response to neutral faces (A) and fearful faces (B) with whole-brain correction at significance level of $P_{FWE} < 0.05$ . Global peaks (visual cortex) highlighted with red arrows.

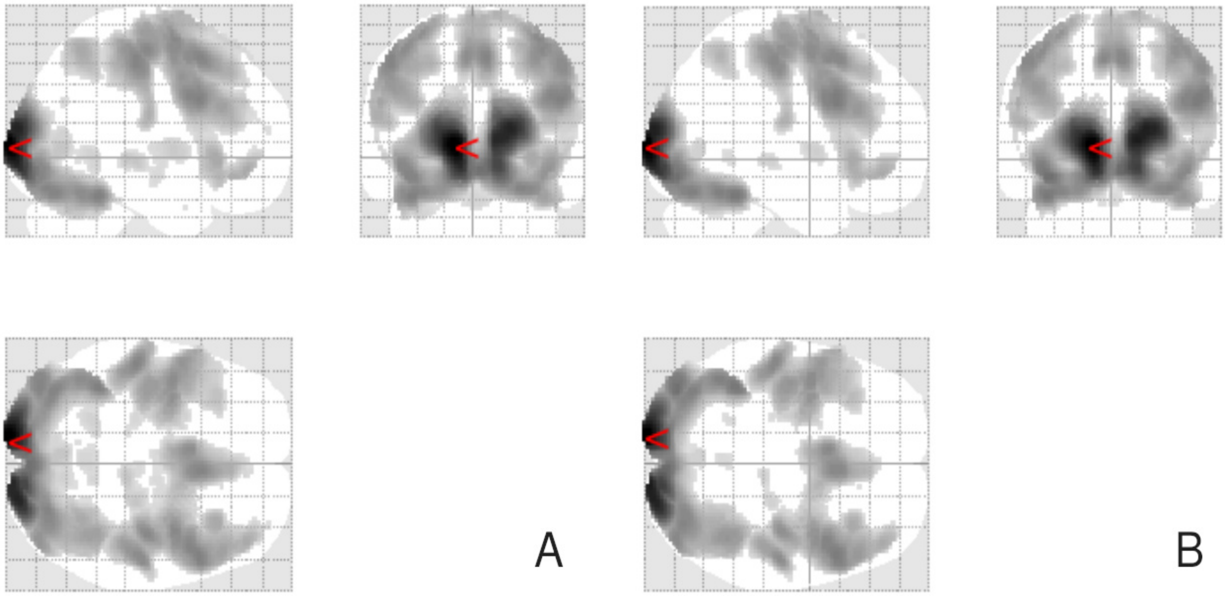

**Figure S4 Increased whole-brain activations to neutral faces in LMDD.** 'Glass' brain illustration of increased whole-brain (cluster-corrected) activation in response to *neutral* faces (Neutral > Baseline contrast) in LMDD compared to controls, without correction for antidepressant medication status (A) and with correction (B). Red arrow at MNI -18 30 30 (A) and MNI -28 10 42 (B).

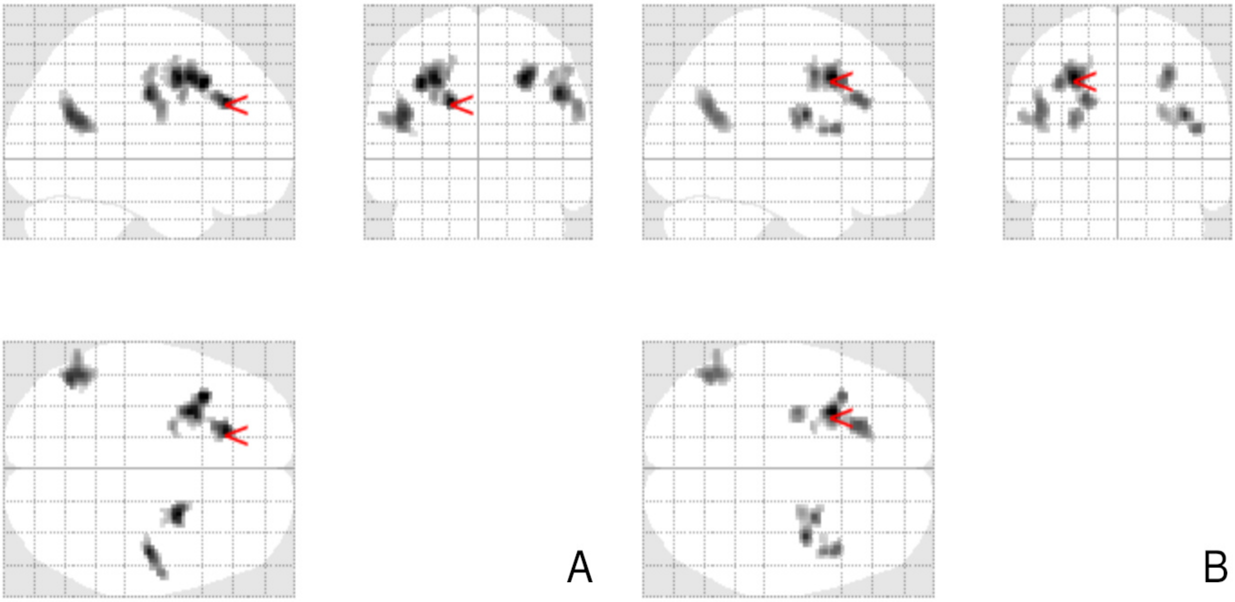


**Figure S5 Increased whole-brain activations to fearful faces in LMDD.** 'Glass' brain illustration of increased whole-brain (cluster-corrected) activation in response to *fearful* faces (Fearful > Baseline contrast) in LMDD compared to controls, without correction for antidepressant medication status (A) and with correction (B). Red arrow at MNI 2 -16 44 (A) and MNI -20 -2 46 (B).

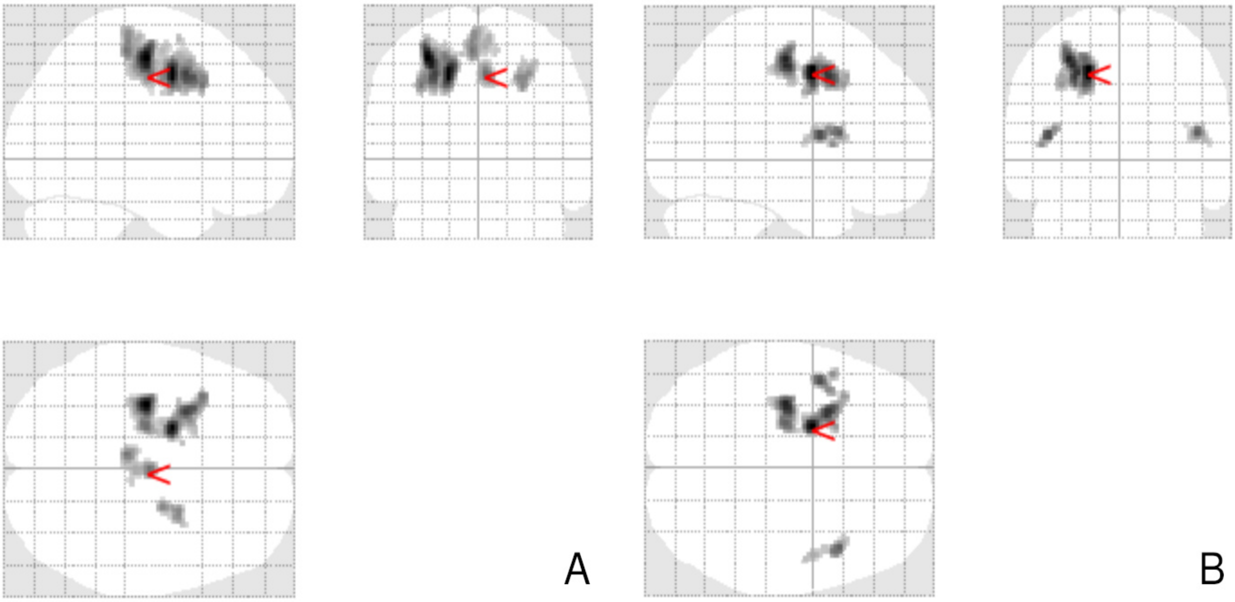

**Figure S6 Increased DLPFC activations to neutral faces in LMDD.** ’Glass’ brain illustration of increased activation with DLPFC SVC in response to *neutral* faces (Neutral > Baseline contrast) in LMDD compared to controls, without correction for antidepressant medication status (A) and with correction (B). Red arrow at MNI -36 18 40 (A) and MNI -30 12 42 (B).

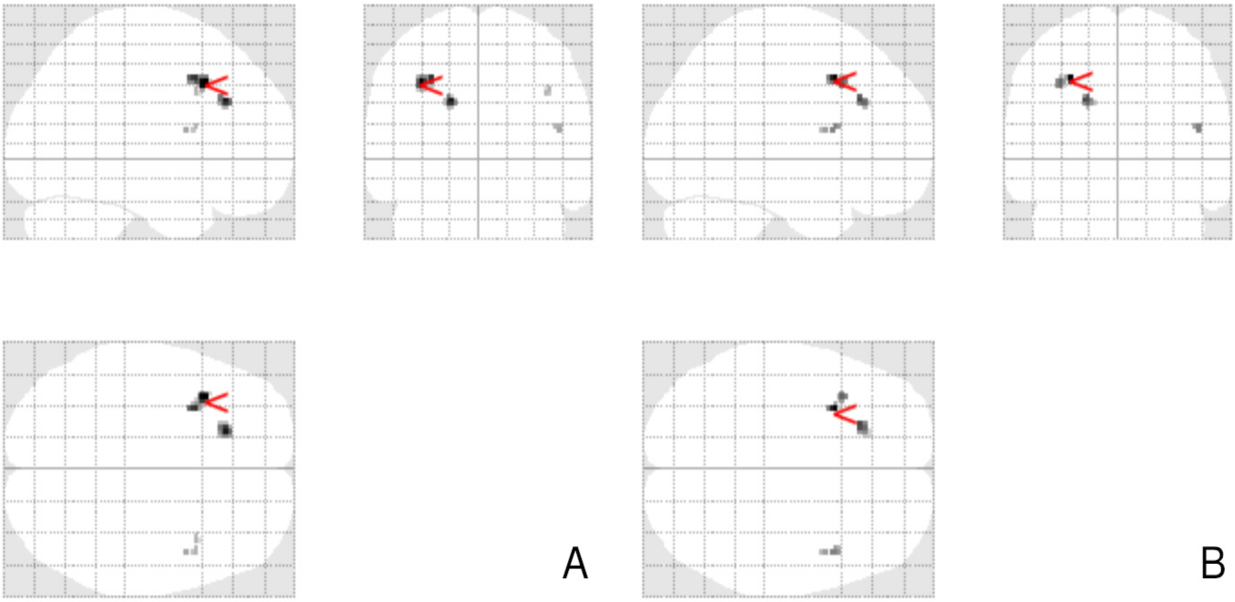


**Figure S7 Increased DLPFC activations to fearful faces in LMDD.** ’Glass’ brain illustration of increased activation with DLPFC SVC in response to *fearful* faces (Neutral > Baseline contrast) in LMDD compared to controls, without correction for antidepressant medication status (A) and with correction (B). Red arrows at MNI -30 8 42 (A) and MNI -30 10 42 (B).

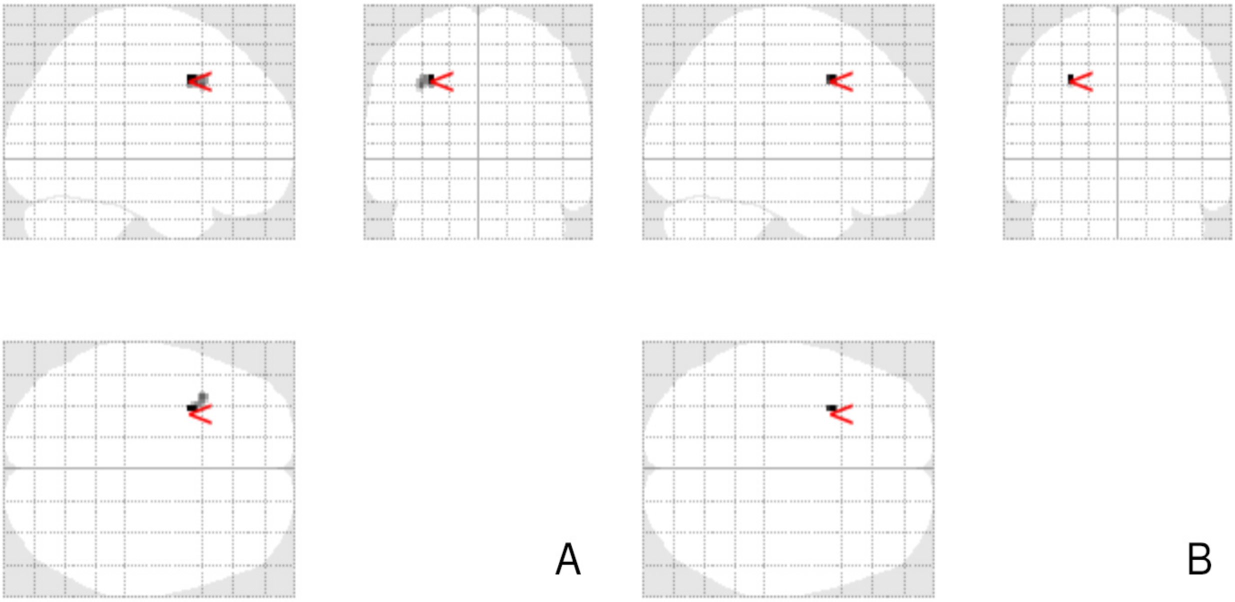
